# A probabilistic graphical model for estimating selection coefficient of nonsynonymous variants from human population sequence data

**DOI:** 10.1101/2023.12.11.23299809

**Authors:** Yige Zhao, Tian Lan, Guojie Zhong, Jake Hagen, Hongbing Pan, Wendy K. Chung, Yufeng Shen

## Abstract

Accurately predicting the effect of missense variants is important in discovering disease risk genes and clinical genetic diagnostics. Commonly used computational methods predict pathogenicity, which does not capture the quantitative impact on fitness in humans. We developed a method, MisFit, to estimate missense fitness effect using a graphical model. MisFit jointly models the effect at a molecular level (*d*) and a population level (selection coefficient, *s*), assuming that in the same gene, missense variants with similar *d* have similar *s*. We trained it by maximizing probability of observed allele counts in 236,017 European individuals. We show that *s* is informative in predicting allele frequency across ancestries and consistent with the fraction of de novo mutations in sites under strong selection. Further, *s* outperforms previous methods in prioritizing de novo missense variants in individuals with neurodevelopmental disorders. In conclusion, MisFit accurately predicts *s* and yields new insights from genomic data.

## Main

Missense variants, which cause single amino acid changes in proteins, are the most common type of variant in protein-coding regions. They are major contributors to genetic risk of developmental disorders^1-3^, cancer, and other diseases. However, as missense variants have a potentially broad range of functional impact but generally a low chance of recurrence, most missense variants identified in cohorts or clinical sequencing have uncertain effect^4-9^. Deep mutational scanning (DMS) assays can assist with interpretation of missense variants^10-31^, but there is limited scalability as different proteins have different and multifaceted functions that require different functional assays. Therefore, computationally predicting the effect of missense variants is important to support the scale required for novel disease gene discovery and interpretation.

Although many methods have been developed to predict variant effects, there is a long-standing ambiguity of the concepts used to describe variant effect. We adopt a set of definitions^32^ to explain the related causes and consequences specifically for different aspects of missense variant effect (Supplementary Fig. 1). At the molecular level, we define the effect (*d*) as change of abundance, localization, or function of a protein. At organism level, a damaging variant (with larger *d*) is defined as pathogenic if it increases the risk of diseases or conditions. Pathogenic variants are often the focus in human genetic studies and clinical testing. Databases like ClinVar^9^ and HGMD^33^ have curated pathogenic variants, which are used as the training labels in supervised methods, such as CADD^34^, REVEL^35^, M-CAP^36^, gMVP^37^, VEST^38^, MetaSVM^39^, MVP^40^ and MPC^41^. Although these methods have proven helpful, they usually suffer from inconsistent performance across genes, since most of the curated pathogenic variants are from only a few thousand genes that are well-established as disease-associated. We suggest that predicted pathogenicity is an uncertain aggregation of variant functional effect, gene risk, and even disease properties. Our knowledge of gene to disease association is incomplete and in fact, identification of new associations is a primary goal of predicting variant effect in genetic studies. Therefore, we seek other metrics for describing missense effects in prediction that can be quantified without knowing gene-disease associations.

One such metric is selection coefficient (*s*) which quantifies the fitness effect of variants in a population. A pathogenic variant is usually subject to negative selection in human populations. Although *s* of a variant depends on the penetrance of the variant to various conditions and the total fitness effect of the conditions, the consequence of *s*, especially of heterozygotes, can be observed in allele frequencies in human populations^42^. It is therefore theoretically feasible to estimate *s* without knowing any traits with which the variant is associated. Biobank-scale genomic sequencing efforts^4-8^ have generated a large number of human population genome sequences that enable estimation on heterozygous selection coefficient of gene-aggregated protein truncating variants (PTVs)^43-46^. However, estimation for missense variants is much more challenging because we cannot reasonably assume all or most of missense variants in one gene have the same selection coefficient. Existing prediction of selection for individual variants^47^ does not directly utilize protein context, and is still based on a very small sample size.

Here we describe a new method, MisFit, to jointly predict molecular effect and human fitness effect of missense variants through a probabilistic graphical model. We aimed to estimate selection coefficient for variants under moderate to strong negative selection. In the model, the molecular effect depends on amino acid change in the protein context, and heterozygous selection coefficient depends on molecular effect of the variant and gene-level importance in selection in human populations. We trained the model using population genome data without pathogenicity labels and evaluated it using deep mutational scan readout data and de novo and inherited variants in developmental disorders.

## Results

### Using Poisson-Inverse-Gaussian distribution to model allele counts in human populations

The distribution of allele counts (*m*) in population sequencing samples is determined by heterozygous selection coefficient (*s*), mutation rate (*v*) and number of chromosomes (*n*). To infer *s*, we first need to model the probability of observed allele counts *p*(*m*|*s*; *v, n*). Allele frequency *q* of a variant at equilibrium state equals 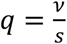, and therefore the allele count *m* follows a Poisson distribution with an expectation 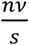. When taking genetic drift into account, the distribution has strong overdispersion. Nei’s model^48^ describes allele count as a Negative Binomial distribution with an additional parameter, effective population size *N*_*e*_. However, as there was exponential growth in recent generations^49,50^, *N*_*e*_ is not a constant, and there is no closed form to describe *p*(*m*|*s*; *v, n*). Here, we used a long-tailed distribution, Inverse-Gaussian (IG) distribution, to approximate the distribution of *q*, which results in a Poisson-Inverse-Gaussian (PIG) distribution of *m*. The parameters associated with the PIG distribution are functions of *s, v, n*, which are optimized prior to MisFit training steps by simulated allele frequencies given dense grids of *s, v* and European effective population size history (Methods). We are mainly interested in those rare variants with relatively large *s*; therefore, we chose to optimize the distribution for the recent exponential population growth. In this way, we were able to easily obtain a tractable gradient to *s* with a time complexity independent to *n*.

To investigate how to approximate allele frequency distribution in a finite and expanding population, we performed a simulation based on a demographic history model of European population^49^. Given *v* and *s*, we sampled each generation by a Wright-Fisher process (Methods). We set the final effective population size to 1.5 million, as it best fits the distribution of observed sample allele counts of rare synonymous C-to-T variants in methylated CpG sites with high roulette mutation rate (*v* > 10^−7^ per generation) (Supplementary Fig. 2). This final population size is smaller than recent work^45,46^ (5 million), which is optimized for all variants with gnomAD mutation rate (with an lower average *v* ∼ 6 × 10^−9^).

We fitted the PIG model parameters (Supplementary Fig. 3) based on simulated variants under different settings of *v* ∈ [10^−9^, 3 × 10^−7^], *s* ∈ [10^−6^, 1]. When *s* is small, random drift makes the distribution of allele counts more resemble a Negative Binomial distribution with small *N*_*e*_.

When *s* is large, the distribution is closer to a Poisson distribution, as these variants are likely to emerge recently when the effective population size is large (Fig. 1a,b). The PIG model fits the simulated results better than other simple distribution models in all ranges (Supplementary Fig. 4).

**Fig. 1.**
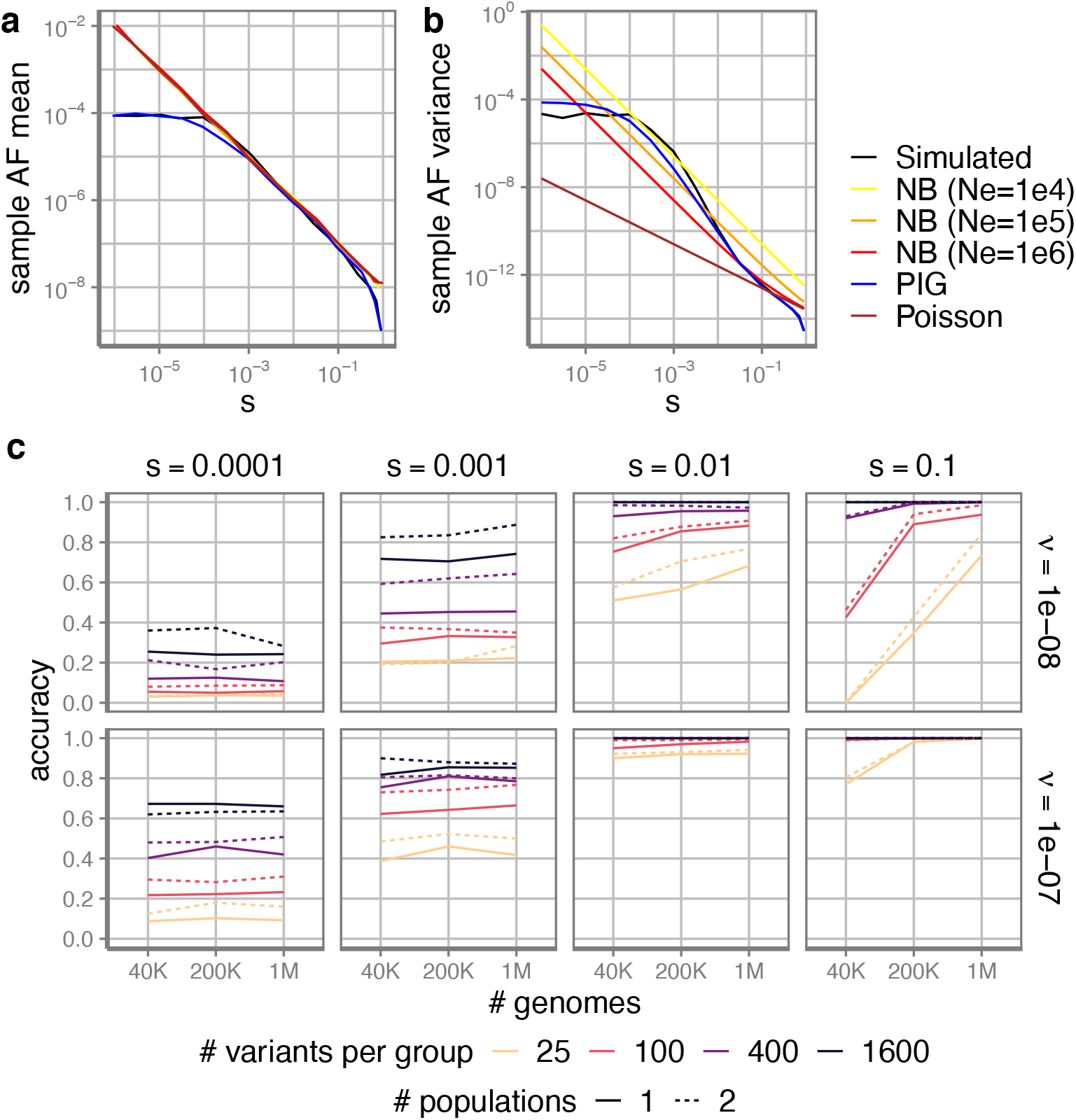
Poisson-Inverse-Gaussian (PIG) distribution with adjusted parameters to approximate allele count distribution. **a** mean and **b** variance of sample allele frequency under different population genetics models, including our PIG model and Negative Binomial model with different effective population size. Diploid sample size is 200K. Mutation rate is 10^−8^. **c** The accuracy of MLE estimation of *s*. Here *s* is a categorical variable of 0.00001, 0.0001, 0.001, 0.01, 0.1, 1. Accuracy is measured by the proportion that the estimated categorical *s* equals the simulated in 400 simulated groups. Each group contains a certain number of variants (x-axis) with same *s*. Solid lines are samples from a single population, while dashed lines are samples from two populations (half of the indicated number for each population).

### Feasibility of estimating selection coefficient for a group of variants

Given the generally low mutation rate at 10^−8^, the highest probability usually lies at 0 count regardless of *s* (Supplementary Fig. 4), so it is nearly impossible to precisely estimate *s* for individual single nucleotide variants only using allele counts. We therefore investigated the feasibility of estimating *s* for a group of variants with similar *s*. We aggregated certain numbers of sites simulated from the same *s* as a group (Fig. 1c). We investigated whether the maximum-likelihood-estimation (MLE) for the whole group based on the PIG model is consistent with the simulation condition.

For deleterious variants of *s* > 0.01 with high mutation rate, the accuracy is high. More variants to aggregate and higher mutation rate always helps with better estimation. The PIG model does not provide good performance for *s* < 10^−4^, because randomly including or excluding a common variant in the group can significantly change the joint likelihood. Notably, increasing sample size in a single population only helps with variants under strong selection (*s* > 0.01) (Fig. 1c, Supplementary Fig. 5). The over-dispersion of allele counts for milder variants mostly comes from the uncertainty of allele frequency (the long-tailed distribution of *q*) due to genetic drift, rather than from sampling (the Poisson distribution given *nq*). Adding samples from another population improves accuracy more than from the same population. Based on the results, we implicitly group missense variants by the degree of damage (*d*) in the same gene in the MisFit model.

### MisFit model structure and training process

We describe the architecture of MisFit in Fig. 2. The degree of damage (*d*) for each single amino acid substitution depends on the protein sequence and structure context. Here we used the embeddings (*x*) from the last layer in the masked protein language model, ESM-2 (650M)^51^ to capture the protein sequence and structure context. We added additional transformer blocks and fully-connected dense layers to generate a distribution of *d* (Supplementary Fig. 6b). Rescaling and normalization of *d* by a gene-level, species-averaged selection strength gives out probabilities of each amino acid at the position. The heterozygous selection coefficient (*s*) depends on *d* and the gene-level selection strength in the human population. Here we modeled *s* in the logit scale as linear to *d*. We set a global prior for the maximum missense selection coefficient for each gene (*s*_*gene*_, the value of *s* when *d* equals 1). (Methods). Finally, probability of generating allele count *n* given *s* is given by the PIG model as previously described.

**Fig. 2.**
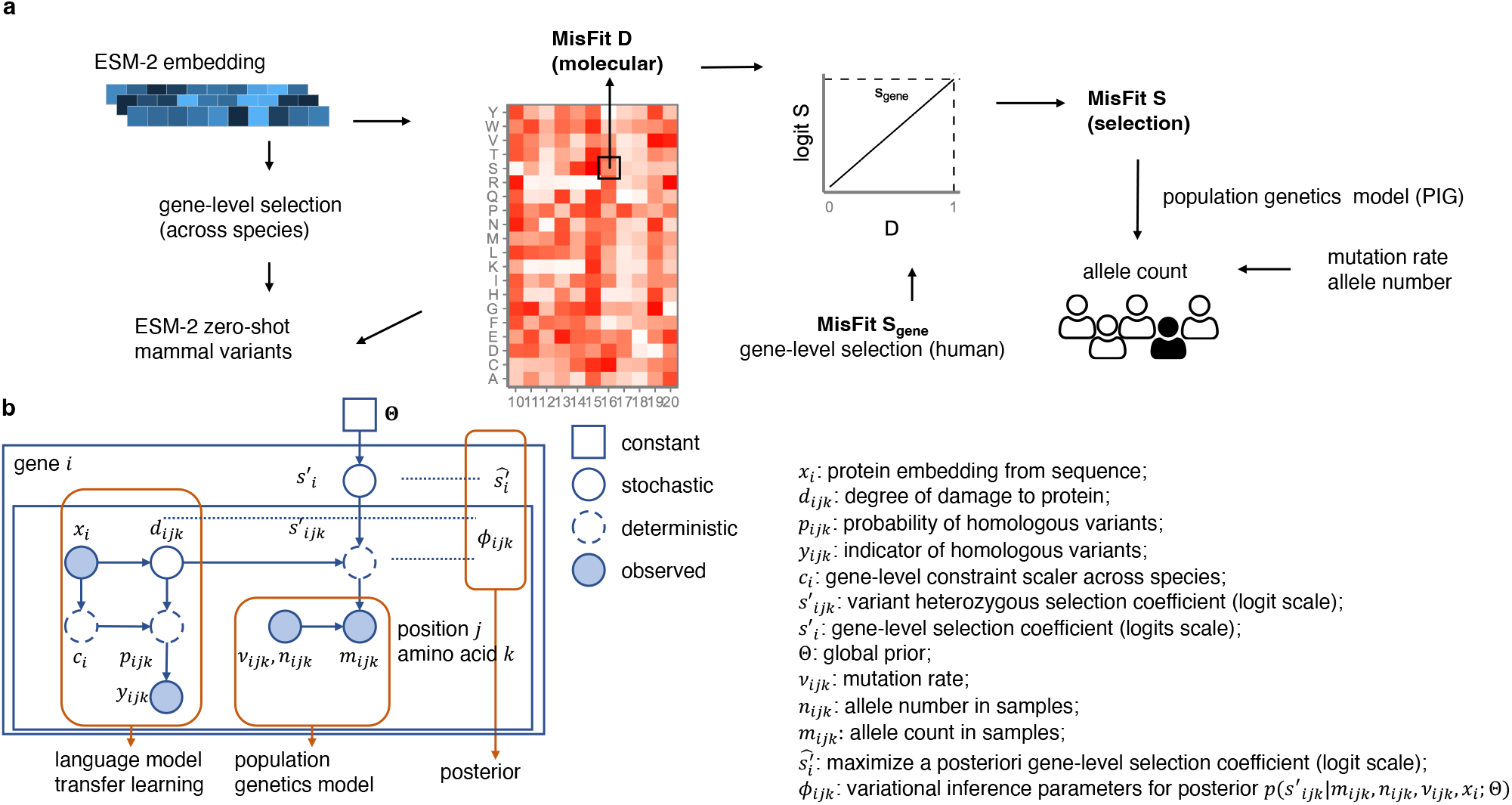
MisFit model for estimating molecular and fitness effect. **a** Overview of MisFit model. MisFit_D is learned from ESM-2 (650M) protein embedding, and generates probability of amino acid in orthologues. Heterozygous *s* is linear to *d* in logit scale, with gene-level maximum from a global prior. MisFit_S is a point estimate of *s* per variant, which maximizes the allele count likelihood in population samples. **b** MisFit model in view of a probabilistic generative process.

In the first stage, we trained the model to estimate parameters in transformer and dense layers (denoted as *NN*^1^ in Methods), to maximize the log likelihood with allele counts, amino acid in orthologues^52,53^, and ESM-2 zero-shot prediction. In other words, we attempt to approximate *p*(*d*|*x*) by maximizing *p*(*m*|*x, v, n*), as this gives out estimation of *d* across genes. During this training stage all possible missense SNVs in 18,708 genes were used, although for most epochs, only a subset of 4,073 constrained genes (missense z score > 2 or gnomAD^6,7^ pLI > 0.5) well covered with both mammal sequence alignment and human population sequence were included, because damaging variants in these genes are expected to be under relatively strong selection, and thus the difference in molecular effect can result in a broad range of selection coefficient for training. Allele counts in 236,017 samples are used in training, including 145,103 UKBB^8^ unrelated European samples and 90,914 gnomAD^7^ Non-Finnish European population samples. Finally, *s*_*gene*_ for all genes were updated by maximum a posteriori (MAP).

In the second stage, with the estimated *d* and *s*_*gene*_, we performed variational inference to approximate the posterior distribution of *s* for each missense SNV. During this stage, *s* is a hidden variable while its prior is regarded as known with the optimized *NN*,, and thus the posterior distribution *p*(*s*|*m*; *x, v, n*) is determined but with no simple analytical form. Sampling-based methods are a solution but time-consuming considering the amount of individual missense variants. Therefore, we treat posterior *s* as functions of *d* and population data, which is modeled by another dense neural network (denoted as *NN*^2^ in Methods) to enable efficient variational inference in one forward-pass. (Supplementary Fig. 6b, Methods)

### Comparison of gene-level constraint

MisFit-estimated *s*_*gene*_ quantifies gene-level selection strength on missense variants. Commonly used metrics for such information include gnomAD missense z score and o/e. Though *s*_*gene*_ for each gene generally correlates well with both metrics (Supplementary Fig. 7), they represent different aspects. Missense z score is effectively the significance level assuming a Poisson distribution from the expected number of variants. Thus, when a gene is short, missense z score tends to have a small absolute value. *s*_*gene*_ and o/e directly represent the degree of constraint, although the uncertainty for short genes might be large.

Previous studies^44-46^ estimated *s* for protein truncating variants (PTVs) in each gene, assuming PTVs in a gene have the same *s*. We compared *s*_*gene*_ with a sampling-based method^45^ for PTVs. PTVs mainly decrease protein levels by nonsense mediated decay. As most of the missense variants are hypomorphs with partial loss of function, *s*_*gene*_ and *s*_*PTV*_ are highly correlated (Fig. 3a). However, some variants can be damaging through mechanisms other than loss of function. We highlighted risk genes with known genetic modes^54^ (Fig. 3b-e, Supplementary Table 1). Autosomal recessive genes are least intolerant of PTVs compared with other genes associated with dominant inheritance. Haploinsufficient genes are under strong selection on PTVs. Genes with dominant negative effects are likely to be under strong selection on missense and PTVs. Notably, for gain-of-function, a subset of genes are only constrained on missense but not on PTVs (Supplementary Fig. 8). For example, several germline missense variants in oncogene KRAS lead to Noonan syndrome by hyperactivation of the protein^55^. The gene-level selection on missense variants is significantly higher than PTVs (*s*_*gene*_ = 0.37, *s*_*PTV*_ = 0.00020).

**Fig. 3.**
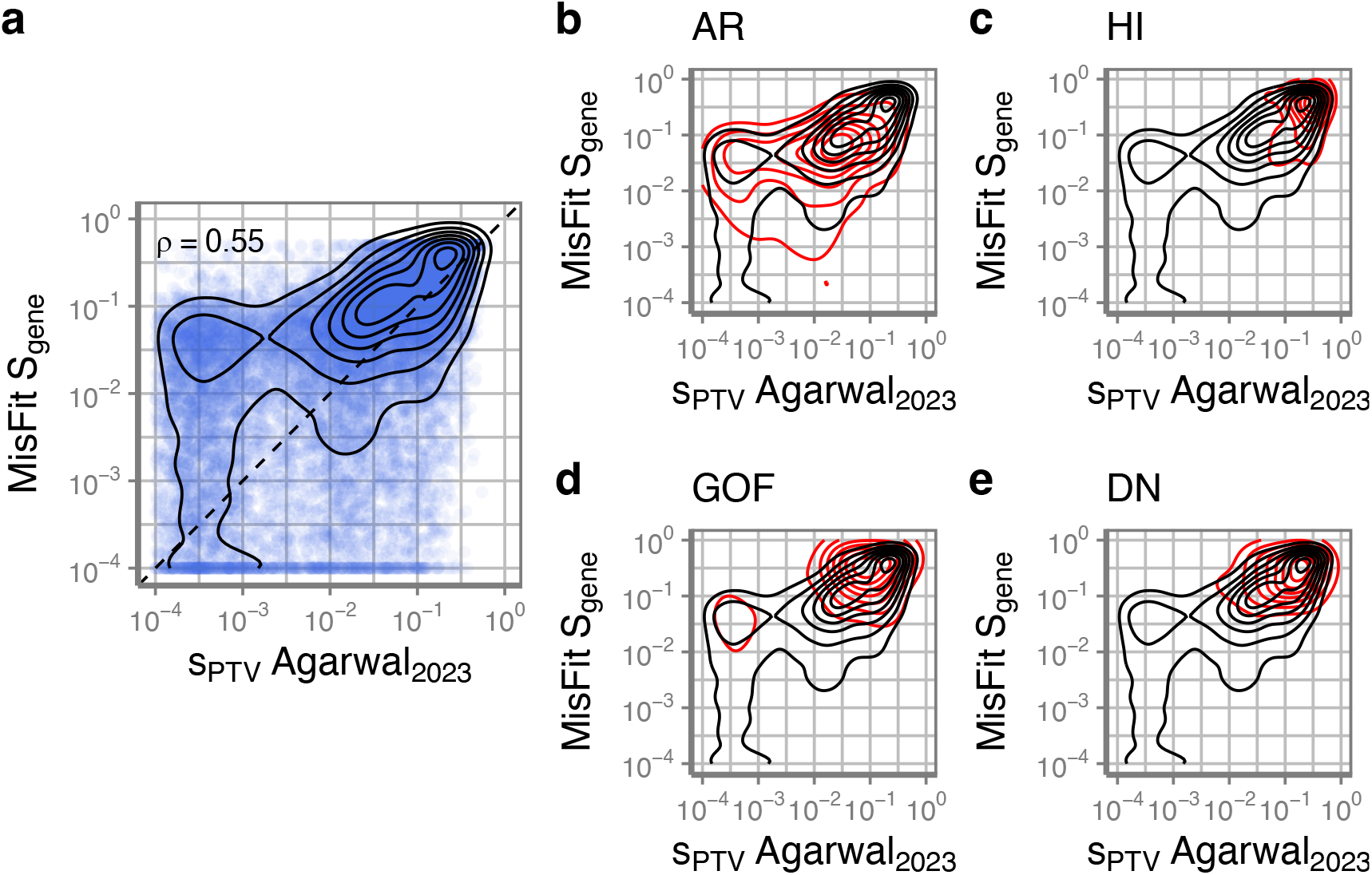
Gene-level missense selection Sgene compared with selection coefficient of protein-truncating variants. **a** the distribution of all genes (black contour) with Pearson correlation coefficient. **b-e** in genes harboring variants of known mechanisms (red contours): **b** autosomal recessive (n = 641) **c** happloinsufficient (n = 66) **d** gain-of-function (n = 69) **e** dominant negative (n = 54).

### MisFit is predictive of allele counts of ultrarare variants in different populations

As MisFit_S is able to predict *s* with amino acid resolution (Supplementary Fig. 9), we asked how informative MisFit_S is to predict allele frequency of rare variants in a population of different ancestries. We extracted 215,138 positions without observed missense variants or with ultra-rare (sample allele frequency < 5 × 10^−6^) missense variants and high mutation rate (*v* > 10^−7^) in 4,073 constrained genes of the training set (UKBB and gnomAD NFE, 236,017 samples, thus allele count ≤ 2 for most highly covered sites). We binned the variants by estimated MisFit_S and analyzed the counts in a second population of a different ancestry, which is gnomAD African/African American (AFR) with 28,872 individuals. Putative variants in these positions would have emerged very recently, and their allele frequencies are relatively independent between the two populations. As expected, the proportion of variants with 0-count in gnomAD AFR samples is positively correlated with MisFit_S (Supplementary Fig. 10a). The opposite trend is observed for the percent of variants with 10 times higher allele frequency in AFR (Supplementary Fig. 10b). To assess which part of the model helps with prediction of *s*, we built several models with fewer and simpler components. In the baseline model (model 0), *s* is estimated from only the mutation rate and allele counts with a global prior. For this chosen set of variants of high mutation rate, allele count is informative as shown in the stepwise curve caused by allele counts of 0, 1, 2 in the training set. However, the difference in absolute value of selection is subtle (Supplementary Fig. 10c). Adding the gene-level selection (model 1) in the model largely improves and smooths the estimation and outputs a wider range of *s*. Using ESM-2 zero-shot score to infer probability of damage (model 2) further helps the prediction, indicated by a greater slope of the monotonic increase, but is not as good as the full MisFit model, which uses the ESM-2 embedding.

### Comparison of selection coefficient with de novo fraction

Next, we evaluate whether MisFit_S approximates heterozygous selection coefficient *s* in absolute scale. We obtained missense de novo (16,876 cases, 5,750 controls) and inherited variants (6,507 cases, 2,992 controls) from an autism spectrum disorder study^1^ (Supplementary Table 2). MisFit_S of de novo variants are significantly higher than inherited variants (Supplementary Fig. 11). We binned the variants based on MisFit_S and normalized the counts as per individual (Fig. 4a). The difference between cases and controls is significant for de novo missense variants for strongly deleterious variants (MisFit_S > 0.01), but is subtle for inherited variants, even if limiting the data to known autism genes (Supplementary Fig. 12).

**Fig. 4.**
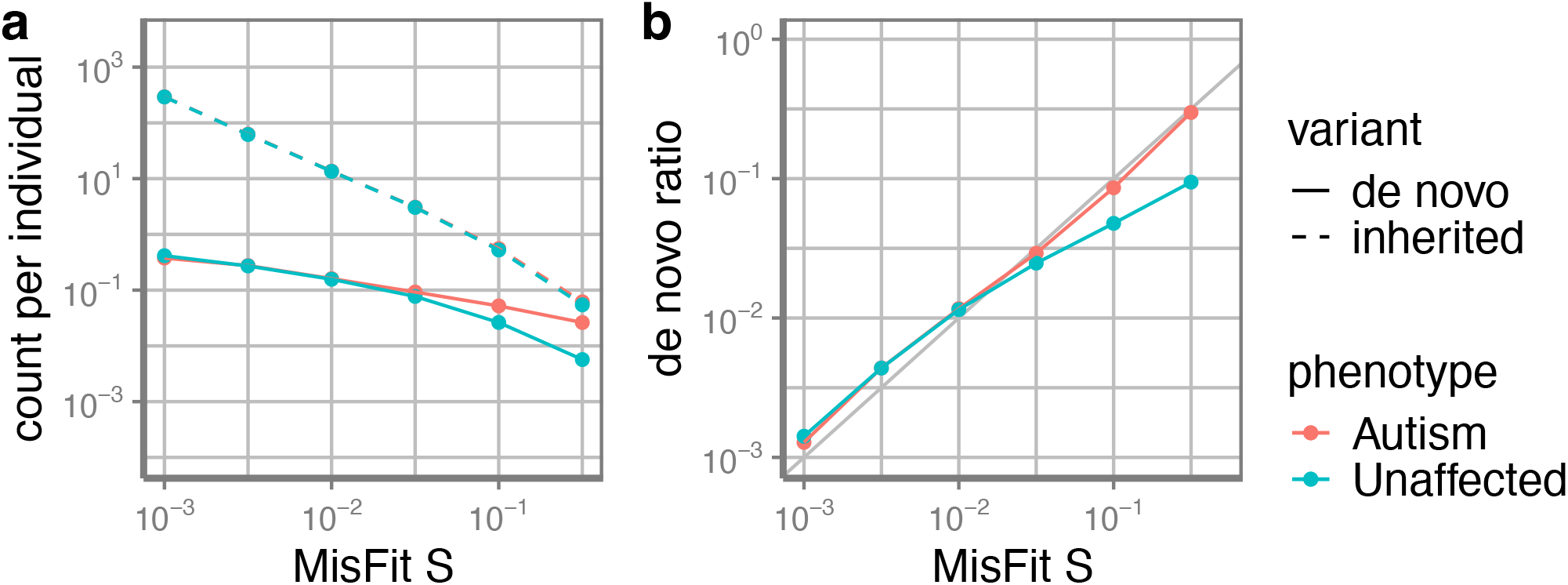
de *novo* or inherited missense variants binned by of MisFit_S. **a** Count of de novo or inherited missense variants. **b** The proportion of *de novo* to all variants in autism dataset. Error bars show 95% confidence intervals.

In a new generation when selection has not occurred, de novo variants are expected to take up a proportion that is equivalent to *s* when *s* is relatively large^44^. We aggregated the variants by their selection coefficient and calculated the fraction from the de novo variants. The de novo ratio in autism cases is consistent with MisFit_S, indicating the accuracy of estimated *s* in absolute scale. In controls, which were unaffected siblings in families ascertained by cases, highly deleterious de novo variants are depleted. (Supplementary Note)

### Analytical utility of selection coefficient for de novo variants in developmental disorders

In addition to the autism data, we obtained de novo variants from studies of neurodevelopmental disorders (NDD, most individuals have global developmental delay or intellectual disability)^56^ (31,565 cases) (Supplementary Table 2). Previous studies^1,3,56^ have shown that a substantial fraction of de novo missense variants in these cases are risk variants for NDD. Autism and NDD are relatively common conditions with early-onset phenotypes.

Autism has a prevalence approaching 0.028^57^, and selection on autism is around 0.7^58^. Thus, highly penetrant risk variants are not likely to be transmitted into the next generation, resulting in a high selection coefficient. As expected, de novo variants in cases have a higher MisFit_D and MisFit_S than controls (Fig. 5). We compared our results with other missense variant effect prediction methods^34-37,51,59-61^. Although there is no ground truth to know which variants actually increase disease risk, we could calculate the enrichment of variants under different thresholds, which is the ratio of number of variants in cases to what is expected in controls (Methods). Among variants ranked in the top 10 percentiles by multiple methods, MisFit_S reached a higher enrichment ratio (Fig. 6) than any other method.

**Fig. 5.**
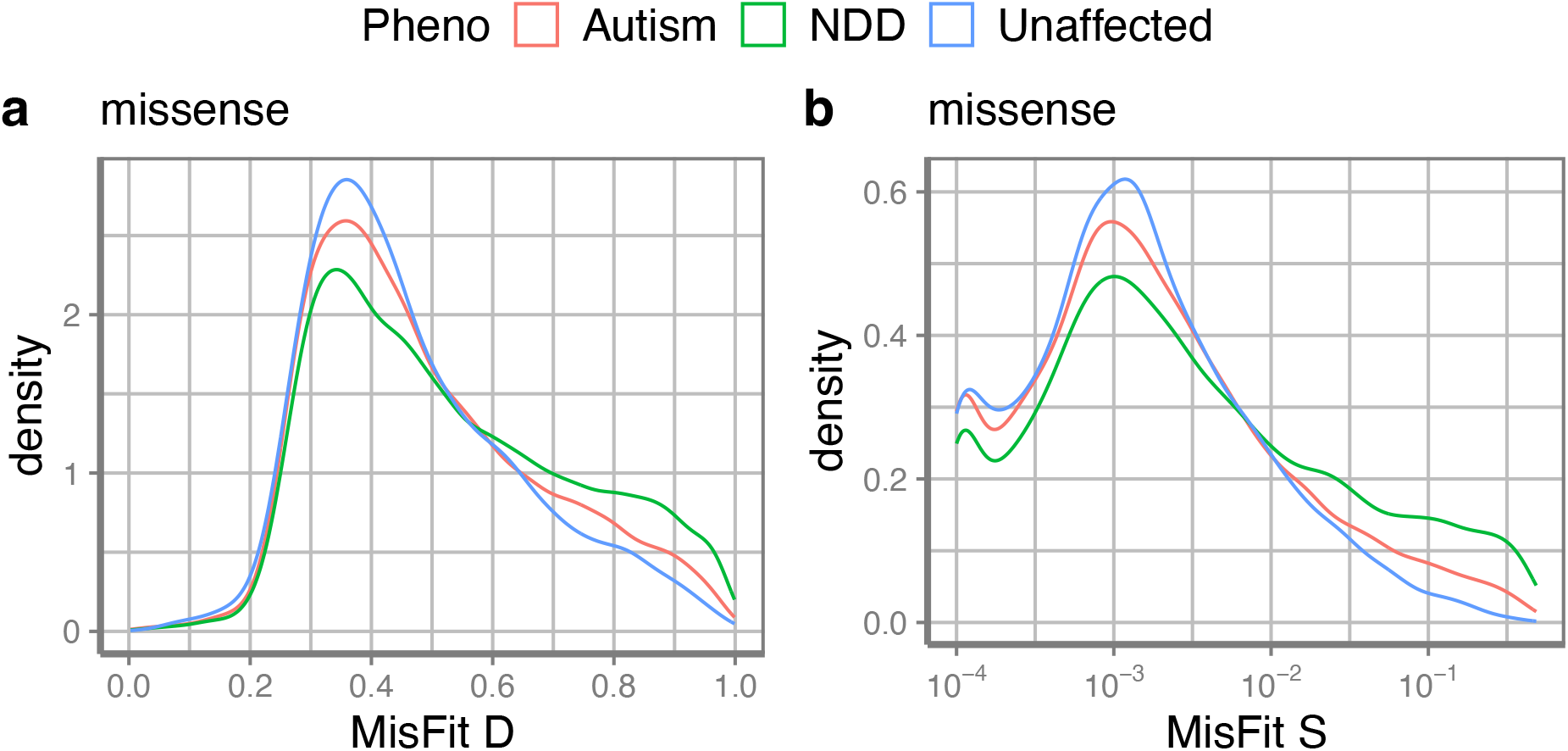
Distribution of MisFit_D and MisFit_S for de novo variants in autism and NDD datasets. Two-sided Kolmogorov-Smirnov tests are used between case and control. a) Missense variants MisFit D (Autism: *p* = 5.5 × 10^−6^; NDD: *p* = 3.2 × 10^−42^); b) Missense variants MisFit S (Autism: *p* = 2.6 × 10^−6^; NDD: *p* = 3.7 × 10^−54^).

**Fig. 6.**
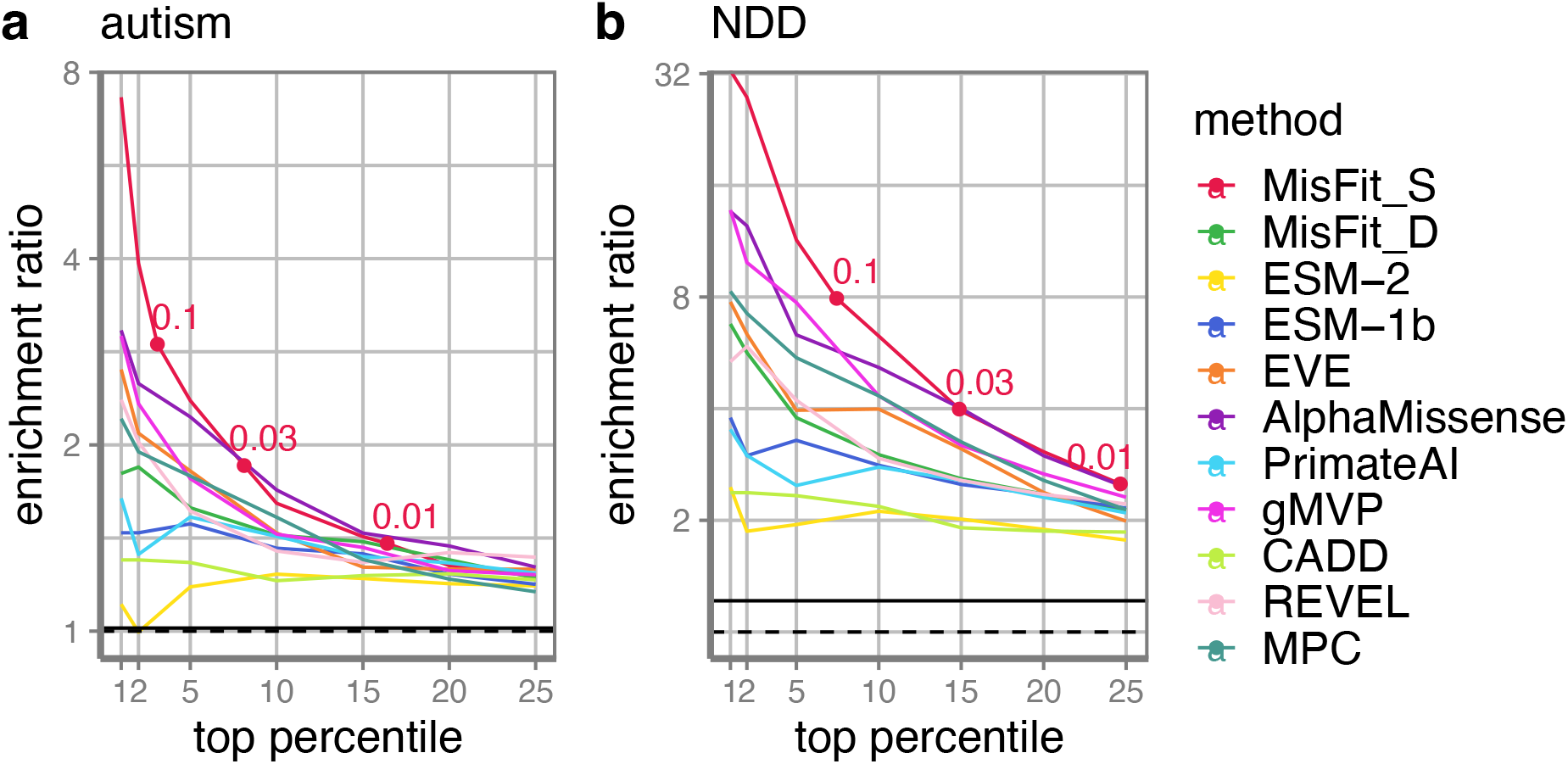
Enrichment ratio of de novo missense variants under different thresholds for multiple prediction methods. Thresholds of MisFit_S are annotated. The solid horizontal line is the overall enrichment ratio including all variants, the dashed line stands for no enrichment.

We then derived the precision-recall-proxy curves (Supplementary Fig. 13, Methods) by the excess number of variants under thresholds. MisFit_S outperforms other methods in high precision range, reaching a precision of 0.67 and 0.87 for autism and NDD, respectively, at MisFit_S = 0.1. The next best methods are AlphaMissense^61^ and gMVP^37^. The estimated precision can serve as weights or informative priors in statistical methods like DeNovoWEST^56^ or extTADA^62-64^ to improve the power in risk gene discovery. The selection coefficients estimated by baseline methods with fewer components are also informative in enrichment of de novo variants but are inferior to MisFit (Supplementary Fig. 14-15).

### MisFit identifies damaging variants consistent with deep mutational scan data

MisFit-estimated *d* (MisFit_D) is about the molecular effect of missense variants, which can be partly measured by deep mutational scanning (DMS) experiments. We compared MisFit_D with published methods^34-37,51,59-61^ on predicting damaging variants in DMS for individual genes.

First, we collected functional readout scores from 32 DMS assays in 26 genes^11-17,19-31^ with 44,100 single amino acid substitutions (Supplementary Table 3). We calculated the Spearman correlation between the functional scores and computational scores (Fig. 7a). MisFit_D has a similar performance with ESM and AlphaMissense.

**Fig. 7.**
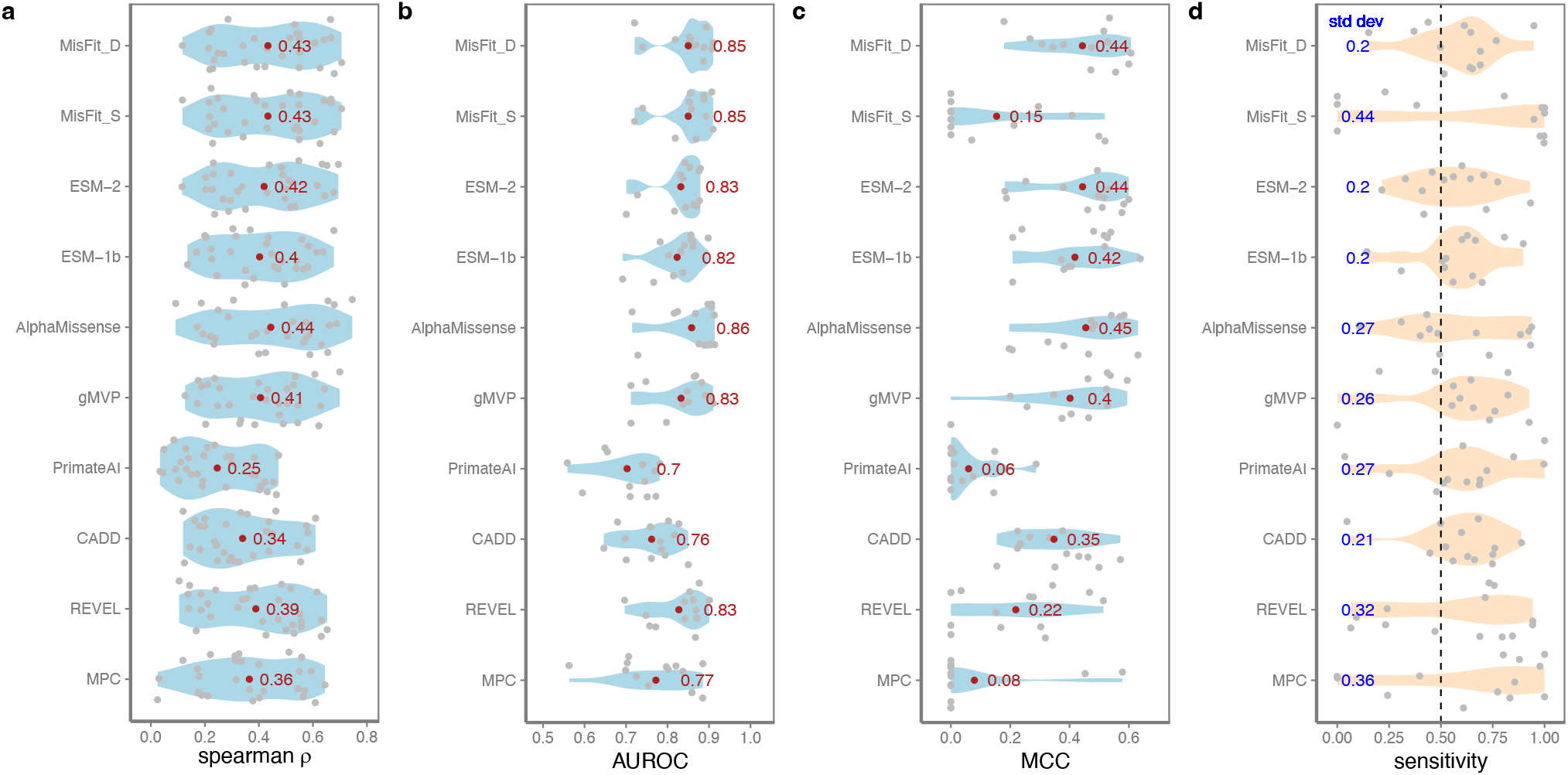
Performance in predicting damaging variants in deep mutational scanning assays and cross-gene consistency. **a** Spearman correlation coefficient of predicted scores with functional scores from deep mutational assays. Mean is annotated in red. **b** AUROC of predicting confidently labeled damaging or benign variants in deep mutational assays. Mean is annotated in red. **c** MCC in each gene with a global threshold that achieves best MCC in the combined dataset. Mean is annotated in red. **d** Sensitivity in different genes when setting a threshold to achieving a global sensitivity of 0.5 (dashed) in the combined dataset. Standard deviation is annotated in blue. For **b-d**, different assays of same gene are combined so that variants with a damaging label in any of the assays will be regarded as damaging.

As raw functional readouts from these experiments could be noisy, we further restricted the sets to variants in 13 genes with DMS annotated binary labels or with a bi-modal functional score distribution (Supplementary Fig. 16). For the latter, we labeled damaging variants by two-component Gaussian Mixture models for each assay independently (Methods, Supplementary Table 4). For genes with multiple DMS assays, we combined these datasets and label a variant as damaging if it is damaging in any one of the assays. The average area under ROC curve (AUROC) for MisFit_D still approaches the state-of-art performance (Fig. 7b).

In some genetic analysis, we often set a heuristic and fixed threshold across all genes when selecting possibly damaging variants. To evaluate the performance under this setting, we combined the DMS assays across genes, and tested the performance in the combined dataset. Since the labels are unbalanced, we define the optimal threshold as that which achieves the highest Matthew’s correlation coefficient (MCC) in the combined dataset. When setting this optimal threshold for classification, we calculated the MCC in each individual gene. MisFit_D remains effective, meaning that the prediction is consistently informative across genes (Fig. 7c). MisFit_D is intended to quantify the degree of damage solely based on variant-level property, and we expect it to be distributed similarly across genes. In contrast, selection coefficient (MisFit_S) is by nature determined by both variant- and gene-level properties and should not have the same range in different genes. Supplementary Fig. 17 shows gene-specific score distribution and optimal threshold.

Finally, we investigated the distribution of sensitivity in different genes (Supplementary Fig. 18). Sensitivity is only related to the damaging variants in the dataset. Deep mutational scanning assays are usually designed to evaluate only one aspect of gene function, so the identified damaging variants could be more reliable, while benign ones may disrupt the protein in some other ways not evaluated by the assays. Under a threshold achieving a global sensitivity of 0.5, MisFit_D has a low variance across genes (Fig. 7d). Overall, unsupervised methods (MisFit, ESM1b and ESM2) have lower variance of sensitivity across genes than supervised methods (gMVP, REVEL, and AlphaMissense).

## Discussion

We developed a probabilistic graphical model, MisFit, to estimate the fitness effect of missense variants using large population sequencing data. Selection coefficient (*s*) is a quantitative measurement of fitness effect that can be informed by allele frequency in human populations, but it is very difficult to estimate for individual variants. MisFit addresses this issue by modeling it as a sigmoid-shaped function of the molecular effect *d* of a variant with a gene-specific prior, and jointly modeling *d* as a non-linear function, approximated by deep neural networks, of its protein sequence context. We trained the model using large sets of population sequencing data without any label of pathogenicity. The estimated *s* is highly correlated with frequency of ultra-rare variants in an independent population. Its value is consistent with theoretical expectation of the proportion of de novo mutations among observed variants in a population.

Previous efforts in estimating gene-specific^6,7,65^ or sub-genic^41,66^ regional constraints of missense variants showed the feasibility of using human population data to identify coding regions that are under strong selection, but these methods are heuristic and do not estimate the effect of individual variants. MisFit is based on population genetics models, representing an improved approach of using large-scale population sequencing datasets for estimating variant effect. Additionally, the effect of a variant at organism and population levels is a combination of how the variant alters the protein and how the protein is involved in key biological processes relevant to human traits and diseases. Two variants with the same degree of damage to protein function may have different effects at the organism and population levels if they occur in different genes. Methods that predict pathogenicity by supervised learning are confounded by gene-level properties, as shown by the large variance of classification accuracy across genes given a fixed threshold evaluated by deep mutational scan data. MisFit’s graphical model is designed to untangle the gene-variant confounding. As a result, MisFit_D has a more consistent scale across genes assessed by mutational scan data; and MisFit_S, as a natural combination of variant and gene properties, has superior performance in prioritizing de novo variants in studies of developmental disorders that have strong negative consequence in fitness.

In a longer timescale across species, negative selection is manifested in conservation among homologous sites. Some unsupervised models, such as ESM^51,67,68^ and EVE^60^, predict amino acid probabilities using representation learning based on massive amounts of protein sequences or multiple sequence alignment of homologous proteins. Those alleles used in training are effectively neutral to become nearly fixed in the corresponding species^69^. When further taking phylogenic history into account, observed sequences are correlated, but the distribution may deviate from the stationary distribution of fitness landscape. Although these models are empirically effective^70,71^, for relatively large *s*, all such deleterious variants are likely to be depleted from the collection of wild-type sequences which are mostly fixed alleles in each species, so theoretically their difference in *s* cannot be easily estimated from MSA alone.

Such resolution in estimating relatively large *s* is especially important for analysis of rare variants in genetic studies of early onset conditions. If we assume an early onset condition is the main trait under selection for a risk variant, then the selection of the variant could be approximated as prevalence × relative risk × selection of the condition. Thus, risk variants in conditions with high prevalence and low fecundity, such as intellectual disability and autism^58^, tend to have large selection coefficient. This explains why MisFit shows superior performance in prioritizing de novo variants in autism or NDD datasets. Additionally, the fitness effects of protein truncating variants and missense variants estimated by MisFit using the same data are directly comparable in a quantitative way. This could improve the power of identifying new risk genes and help characterize genetic etiology of human diseases.

We used the embeddings from a protein language model (ESM-2) to represent protein sequence context as the input for the non-linear function that predicts the effect at molecular level. ESM-2 embeddings implicitly capture protein structure information^51^. Explicitly representing protein structure features as input^61,53^ may improve prediction by better capturing residue interactions and critical sites.

Finally, based on the simulation results, the accuracy of MisFit in estimating mildly deleterious variants (*s* < 0.001) is limited. Random drift of these variants causes significant dispersion of allele frequency. Merely increasing the sample size of the same population does not help with the estimation. On the other hand, including diverse populations with different continental ancestry in training would improve the accuracy, as ancestral effective population size increases and variance from genetic drift decreases. We expect that the sample size of non-European individuals with genome sequencing will increase substantially in the near future from ongoing efforts such as gnomAD^7^, All of Us^5^, GenomeAsia^72^, and the Three Million African Genomes project^73^. We will be able to use these data to improve estimation of fitness effect of variants under moderate selection in the future.

## Methods

### Simulation based on European effective population size history

We simulated the distribution of allele frequency based on the history of effective population size of European population for 10,000 generations. We obtained the European effective population size history from the Schiffels and Durbin model^49^. We smoothened the data by setting a growth rate for each period and adjusted the final effective population size to 1.5 million, which is most consistent with distribution of observed allele counts of rare synonymous variants with high roulette mutation rate (*v* > 10^−7^). We assume no linkage and the same background mutation rate (using the average mutation rate), no positive selection effect, and each locus obtains one type of mutation at most. We simulated the evolution of alleles with dense grids of mutation rates *v* ∈ [10^−9^, 3 × 10^−7^], and selection coefficients *s* ∈ [10^−6^, 1]. For a given mutation rate and selection coefficient, we simulated 100,000 independent sites. The simulation follows the Wright-Fisher process considering mutation, drift and selection. We set a backward mutation rate of *v*_1_ = 10^−8^. Suppose the effective population size at *t*^*th*^ generation is *N*_2_, we have

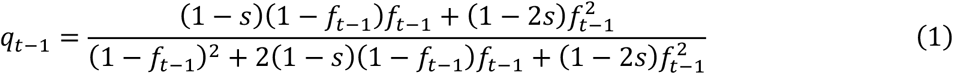

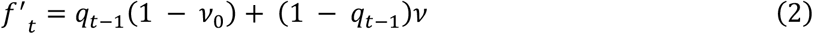

*f*_*t*−1_ and *q*_*t*−1_ are the pre-selection and post-selection allele frequency in the previous generation, and *f′*_2_ is allele frequency in zygotes after introducing new mutations. Here, 2*s* (clipped at 1 if *s* > 0.5) is homozygous selection coefficient by fixing a dominance factor of 0.5. Then we sample population allele counts in the new generation by a binomial distribution:

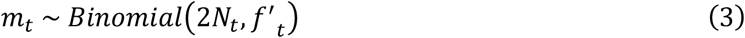

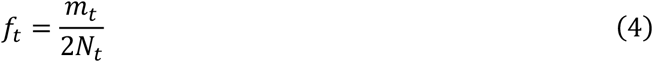

In the latest generation, sample allele counts *m* within sample allele number *n* drawn from population could be regarded as a Hypergeometric distribution.

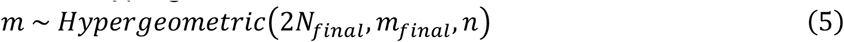

As we have *m*_*final*_ ∼ *Binomial*(_*final*_, *f′*_*final*_), this is equivalent to

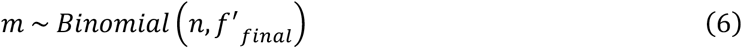

Considering the age of sequencing samples (UK Biobank and gnomAD) are relatively old, the observed alleles are already subject to selection. We therefore used the adjusted post-selection allele frequency for training the model.

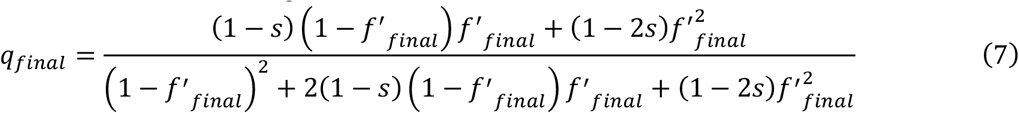

To investigate how a second population with a different genetic ancestry can help with estimation, we simulated a pseudo-population with the same European population size history. Here, *q* is kept same for both populations at the beginning, and then evolves independently for the recent *N*_*r*_ generations. We set *N*_*r*_ to be 2,000 based on the split time of European and Africa population. In this way, the final *q* in two populations are partially correlated.

### Modeling allele counts

Assuming infinite effective population size, allele frequency *q* at equilibrium state is deterministic given mutation rate *v* and heterozygous selection coefficient *s*.

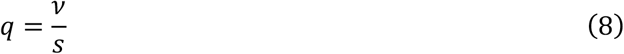

Therefore, the allele count *m* in samples with allele number *n* follows a Poisson distribution:

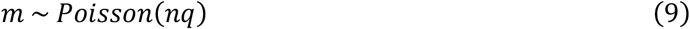

Although the formula gives us an overview of the relationship between expected *m* and *s*, there is a substantial overdispersion of *m* caused by random drift effect. Taking random drift into account, Nei’s model^48^ describes *q* as a Gamma distribution.

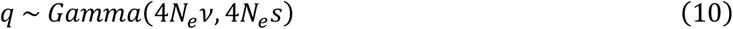

*N*_*e*_ is the effective population size. Then we have a Negative Binomial distribution for *m*.

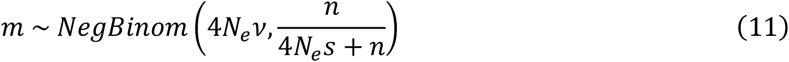

However, the real *N*_*e*_ is not constant. There has been exponential population growth in all major continental populations. We used an Inverse Gaussian model with adjusted parameters *μ*_*IG*_, *λ*_*IG*_ to describe the distribution of allele frequency. Inverse Gaussian distribution can model a very long tail while keeping the probability density at 0 to be 0 (In contrast, Gamma distribution may give out infinity density at 0). More importantly, the likelihood function *p*(*m*|*s*; *v, n*) should have a tractable gradient to *s*. Then *m* follows a Poisson Inverse Gaussian (PIG) distribution:

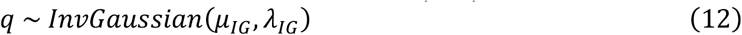

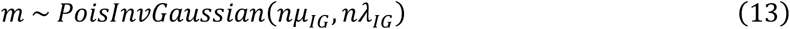

*μ*_*IG*_ and *λ*_*IG*_ are Inverse Gaussian mean and shape respectively. For each setting of *v, s*, we used the simulated allele frequency *q*_*sim*_ to estimate *μ*_*IG*_ = *mean*(*q*_*sim*_),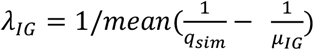. Then we fit functions *μ*_*IG*_ = *f*_1_(*v, s*) and *λ*_*IG*_ = *f*_2_(*v, s*). Specifically, log *μ*_*IG*_ is a softminus over s^*′*^ = logit(*s*) and linear over log *v*, while log *λ*_*IG*_ is quadratic to log *v* (Supplementary Fig. 3). The likelihood of PIG distribution is calculated by Bessel function of second kind.

### Data used in training and testing

#### Proteins and variants

We limit the gene set to 18,708 protein-coding genes. One protein sequence is selected per gene, based on the following order: 1. Uniprot canonical isoform; 2. Corresponding to the transcript of ‘MANE select’; 3. Corresponding to Ensembl canonical transcript (usually the longest). Among them, 18,605 have available population sequencing data for missense variants and 16,623 for protein-truncating variants. All possible single nucleotide variants in the coding region +-2 bp of the selected transcripts are annotated. For protein-truncating variants, ‘stop-gained’, ‘splice_donor’ and ‘splice_acceptor’ variants are further annotated by LOFTEE^6^, and only high-confidence (HC) ones are used in training or genetic analysis.

#### Population sequence data

We used the allele counts from UKBB unrelated European population (145,103 exomes from November 2020 release) and gnomAD Non-Finnish European population (56,885 exomes of v2.1.1 plus 34,029 genomes of v3.1.2) sequencing data. These datasets only contain observed variants in these individuals, but the vast majority of possible but not yet observed variants are also important for estimation. We include all possible missense variants that could result from a single nucleotide substitution. We set the allele number (sample size) for positions without observed variants by the allele number of the nearest position with observed variant in the same exon, to account for sequencing depth variation, and the allele count of these non-observed variants to 0. Same was done for gnomAD African / African American population (8,128 exomes plus 20,744 genomes) in analysis. Variants that do not pass RF, InbreedingCoeff or AS_VQSR filtering, or are located in low-complexity-region (annotated by gnomAD), are excluded in training and analysis.

Site-specific mutation rate was mainly obtained from roulette mutation rates. Variants on sex chromosomes do not have available roulette mutation rates, so we used gnomAD^6^ mutation rates based on 3-mer context and methylation level, and calibrated them to an average of 10^−8^ in consistent with roulette. During training, mutation rate and allele count are added across all single nucleotide variants that lead to the same amino acid change.

#### Protein sequence embeddings

Protein sequence embeddings are extracted from the last layer of ESM-2 (650M) model for each 600 AA length fragments (overlapping 200 AA if longer than 600 AA). The zero-shot prediction of ESM-2 comes from logits value in the last layer of ESM-2 and further renormalized to 20 amino acids excluding other tokens.

#### Mammalian homologues

Homologous variants used in training include: a. 21.8 million alternative amino acids in multiple sequence alignment in 465 mammals from Zoonomia Project^52^; b. 2.9 million alternative amino acids in 233 primate species from primateAI-3D^53^.

#### Deep mutational scanning assays

We selected 32 deep mutational scanning assays from literature (Supplementary Table 3). Several experiments provide classification of damaging or benign variants in the publications. For the remaining experiments, we model the functional scores (usually as log enrichment or depletion) by a two-component Gaussian mixture for each experiment. Amino acid substitutions with probability of damaging > 0.75 are defined as damaging and that < 0.25 are defined as benign. We selected experiments with bimodal score distribution, of which the confident damaging + benign variants make up more than 90% of all variants. In total, 13 genes with damaging / benign labels were selected for evaluating AUROC and MCC (Supplementary Table 4). If there are multiple assays for the same gene (CYP2C9, PTEN, VKORC1), we took the union of damaging labels as positives.

#### MisFit model architecture and parameters in view of a probabilistic graphical model

For a gene *i*, the maximum heterozygous selection coefficient for missense variants is denoted as *s*_*i*_. In our model settings, *s*_*i*_ is transformed into logit scale 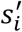 to facilitate numeric computation. For each variant *k* at position *j, d*_*ijk*_ is assumed to be a random variable of logit-normal distribution.

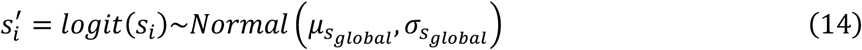

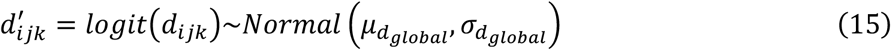

Note that in the full MisFit model, distribution of *d*^*′*^ is learned by neural networks (*NN*^1^) as functions of the protein embeddings *x*_*i*_.

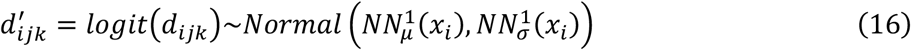

For variant-level heterozygous selection coefficient 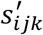, we assume it’s linear to *d*_*ijk*_ (ranging from 0 to 1), where the minimum is set to *logit*(10^−4^) and the maximum is *s*_*i*_ *′*.

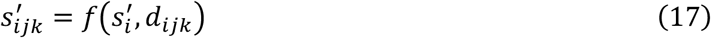

The purpose of our model is to approximate *p*(*d*_*ijk*_ | *x*_*i*_) and *p*(*s*_*ijk*_ | *x*_*i*_, *v*_*ijk*_, *m*_*ijk*_, *n*_*ijk*_), by two parts of MisFit model, *NN*^1^ (functions of only *x*) and *NN*^2^ (functions of *NN*_1_(*x*), *m, n, v*), respectively. MisFit_D and MisFit_S are the point estimates from these two probabilities, which will be described in the next section.

### Model training

The MisFit model contains 4.4M parameters in total. Training of MisFit involves several stages.

In stage 0, before the construction of full MisFit model, we trained a baseline model (corresponding to model 1 in main text and Supplementary Figs. 10, 14, 15). We estimated 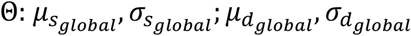 by maximizing 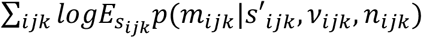, where *s′*_*ijk*_ is calculated from samples of *d*_*ijk*_ given the global priors (Eq. 15, 16). Then we set 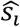 as maximize a posteriori estimation of *s*_*i*_.

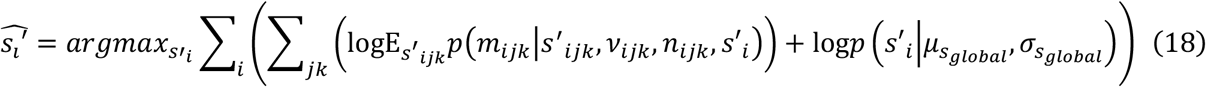

This value of 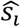 is then used to initialize the main MisFit model.

In stage 1, we aimed to optimize the parameters in *NN*^1^ which connects *x*_*i*_ to *d*_*ijk*_ (Eq. 16). In brief, we would like to estimate

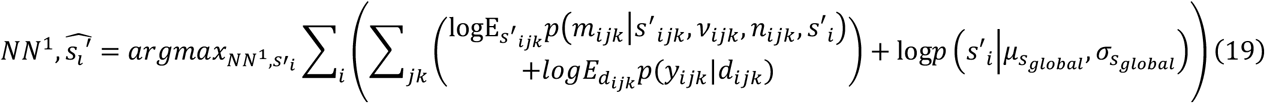

where 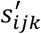 is sampled from Eq. 16,17. This stage involves in several periods. Here *y*_*ijk*_ is a Bernoulli variable denoting where such amino acid change exists in wildtype homologue sequences, where the Bernoulli logit is simply *d*_*ijk*_ transformed by scaler *c*_*i*_. First, we trained *NN*^1^ using all missense variants 13,406 genes well covered with both mammal sequence alignment and human population data for 30 epochs with initial learning rate as 0.001.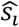 was temporarily set as the value in stage 0. Then we trained *NN*^1^and 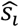 as well on 4,073 constrained genes (gnomAD^6,7^ missense z score > 2 or pLI > 0.5) for 50 epochs with initial learning rate 0.0005. Finally, we kept *NN*^1^ and further inferred 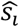 for all genes for 30 epochs.

In stage 2, we did variational inference on the posterior distribution.

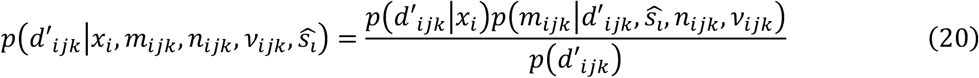

Here, distribution of *s′*_*i*_ is simply represented by its point estimate 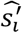. We used a Normal distribution 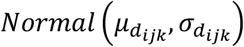 as variational family to approximate this distribution. In order to retrieve 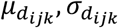 in one forward pass, they are modeled as functions in a second neural network (corresponding to dense layers *NN*^2^ in stage 2 in Supplementary Fig. 6).

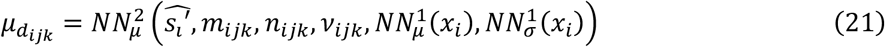

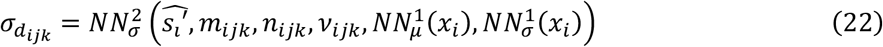

Then like a variational autoencoder, optimizing the evidence lower bound (ELBO) is equivalent to maximizing

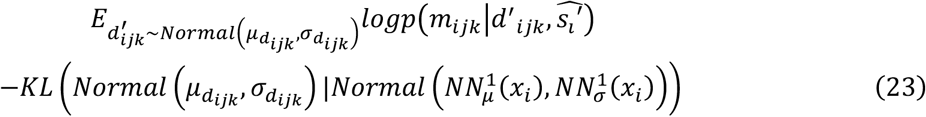

KL() represents Kullback-Leibler Divergence.

During this stage, 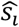 and parameters in *NN*^1^ were fixed and only *NN*^2^ is updated for 20 epochs with initial learning rate of 0.001.

MisFit scores are specifically defined as follows:

MisFit_D: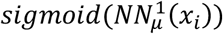, the mean of 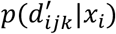 transformed back to original scale.

MisFit_Sgene: 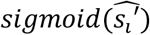, the MAP estimation of 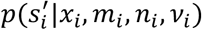 transformed back to original scale.

MisFit_S: 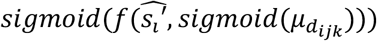, the derived 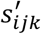 when using the point estimate of 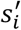, and the point estimate of posterior mean of *d*_*ijk*_ given by Eq. 21.

In our model, the random variable *s* are all represented in logit scale, and our point estimate of *s* is also inferred in logit scale then transformed back to the original scale. This eases the calculation and potentially limits the systematic bias (Supplementary Note).

The main training stage 1 takes around 10 hours on 2 NVIDIA A40 GPUs.

### Enrichment of de novo variants and estimated precision-recall

De novo missense variants in 4 previous genetic studies are used for analysis (Supplementary Table 2). Given a score threshold (to enrich disease risk variants), the number of selected variants is *m*, and *m*_1_ in cases and controls respectively. These numbers are normalized by number of synonymous variants 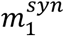 and 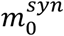 to calculate the enrichment ratio.

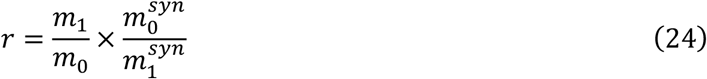

Sensitivity (recall approximate) is estimated by the total number of excess of variants comparing cases and control.

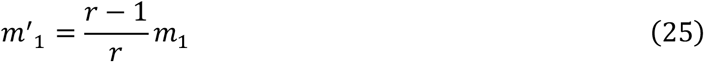

Precision is estimated by

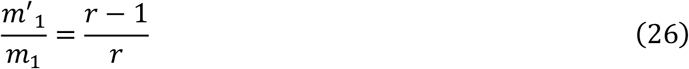

## Data availability

The code for model training and analysis could be found at https://github.com/ShenLab/MisFit. Prediction results, including MisFit_D, MisFit_S for missense variants, and MisFit *S*_*gene*_ for each gene, are available at <u>Dropbox</u>.

## Acknowledgements

We thank gnomAD and UK biobank investigators for making genomic data available. We thank the SPARK study and especially the thousands of families participating in SPARK. This work is supported by NIH grants (R35GM149527, R01GM120609, and P50HD109879), Simons Foundation (SFARI #1019623), and Columbia Precision Medicine Pilot grants program. We thank Itsik Pe’er, Molly Przeworski, Mohammed AlQuraishi, Ben Hu, Adi Garg, Alan Tian, Haicang Zhang, Na Zhu, Chang Shu, and Lu Qiao for helpful discussions.

