## Supplementary Notes and Figures for "A probabilistic graphical model for estimating selection coefficient of nonsynonymous variants from human population sequence data"

Zhao et al

#### Table of Contents

***Supplementary Notes*** ... ***3***

**Poisson-Inverse-Gaussian model for approximating allele count likelihood** ... **3**

**MisFit estimates selection coefficient with amino acid resolution** ... **3**

**Fraction of *de novo* mutations among all variants with a selection coefficient in populations ascertained by a clinical condition** ... **3**

**MisFit performance compared with EVE** ... **7**

**MisFit model on predicting ClinVar variants** ... **8**

**Point estimate for selection coefficient** ... **8**

**Improve estimation of selection by using genomes from different populations** ... **9**

***Supplementary Figures*** ... ***10***

**Supplementary Fig 1 Missense variant effect from different aspects** ... **10**

**Supplementary Fig 2 Adjusted European final effective population size in simulation** ... **11**

**Supplementary Fig 3 Parameters of Poisson-Inverse-Gaussian model** ... **12**

**Supplementary Fig 4 Distribution of sample allele frequency under different population genetics model** ... **13**

**Supplementary Fig 5 Evaluation of MLE estimation of $s$** ... **14**

**Supplementary Fig 6 Overview of MisFit model** ... **15**

**Supplementary Fig 7 MisFit estimated gene-level missense selection correlates with gnomAD missense z score or o/e** ... **16**

**Supplementary Fig 8 Distribution of gene-level selection for genes with known gain-of-function mechanism.** ... **17**

**Supplementary Fig 9 Predicted selection coefficient for each amino acid substitution in PTEN** ... **18**

### Supplementary Notes

##### Poisson-Inverse-Gaussian model for approximating allele count likelihood

In MisFit, the probability of observed allele counts given a selection coefficient in samples is described by a Poisson-Inverse-Gaussian distribution optimizing recent population growth. The choice of Inverse-Gaussian for population allele frequency is heuristic, but it takes account of the long tail, 0 density at 0 and conjugacy to Poisson distribution. In comparison, Nei's model uses a Gamma distribution to approximate the allele frequency, and the density at 0 would be infinity when optimized for a small population size, which leads to a distorted distribution of the resulting Negative Binomial distribution of allele counts. Recent work enables efficient multiplication of the transition matrix of the Discrete-Time Wright-Fisher model[1] to better picture the likelihood, but the PIG model is much simpler with just a few parameters and can be applied to any choice of mutation rate and selection coefficient in a continuous range, which is more applicable to the scale of missense variants. We showed that our method is accurate in estimating relatively strong selection ($s > 0.01$) by simulations.

##### MisFit estimates selection coefficient with amino acid resolution

The main output of MisFit is estimated selection coefficient of an individual missense variant, MisFit_S. We show an example of the phosphatase tensin-type domain in the gene *PTEN* (Supplementary Fig. 9). *PTEN* is a well-known disease risk gene, depleted of missense variants (missense z-score 3.49, o/e = 0.33) and protein truncating variants (pLI 0.26, o/e=0.24). MisFit_S follows secondary-structure patterns, and correlates well with conservation. None of the gnomAD[2] constraint metrics can describe the selection of these sub-gene features. The regional constraint[2], which is calculated by o/e for each 1kb genomic region, provides limited information and low resolution.

##### Fraction of *de novo* mutations among all variants with a selection coefficient in populations ascertained by a clinical condition

Under equilibrium or in a new generation when selection has not occurred, among all observed variants under moderate to strong negative selection in a population, *de novo* mutations are expected to take up a proportion ($\boldsymbol{F}$) equal to the selection coefficient of the variant[3]. Here we provide a proof that this equality still holds for populations ascertained by a clinical condition.

Denote $A$ as the alternative ("mutant") allele and $a$ as the reference ("wildtype") allele of a variant $V$, and the selection coefficient of $A$ as $\boldsymbol{s}$. If $A$ is not associated with the ascertainment condition, then the populations ascertained based on the condition is similar to general population regarding the frequency and proportion of de novos of $A$ ($\boldsymbol{F} = \boldsymbol{s}$ ).

Now we consider a clinical condition with population prevalence $\boldsymbol{\tau}$ ($\tau < 5\%$), assuming that $A$ is associated to the condition, and that the condition is the only condition through which $A$ is under moderate or strong negative selection. As the variant is rare, homozygous genotypes ($AA$) are not considered, and in a proband-parents family at most one parent harboring the heterozygous genotype ($Aa$).

Denote the equilibrium allele frequency of $A$ in a new generation (before selection) as $q$, mutation rate per allele as $\boldsymbol{\nu}$, the selection of the condition as $\boldsymbol{s_0}$, and the penetrance of the condition as $p$. By definition:

$$\boldsymbol{p} = \boldsymbol{P}(\text{Affected}|\boldsymbol{Aa}) \tag{1}$$

In a population with a constant $n$ individuals, there are about $2nq$ individuals with genotype $Aa$, among which $2pnq$ are affected. As a result of the clinical condition, $2pnqs_0$ are negatively selected. Among all individuals with $aa$ genotypes, the number of affected is $2n(1-q)\tau$. Since $\tau$ and $q$ are both small, the impact on variant $V$ due to negative selection on the condition from other factors is negligible. Therefore, by definition, selection coefficient of allele $A$ is:

$$s \approx \frac{2pnqs_0}{2nq} = ps_0 \tag{2}$$

We replace $s_0$ with $s/p$, and the genotypic composition of post-selected individuals, denoted as generation G1 (they are potential parents of the next generation G2) is:

$$P_{G1}(Aa) \approx 2(1-s)q \tag{3}$$

$$P_{G1}(aa) \approx 1 - \ 2(1-s)q \tag{4}$$

The probability that one of the G1 parent is heterozygous is:

$$P(\text{G1: } Aa \text{ and } aa) = 2P_{G1}(Aa)P_{G1}(aa) \approx 4(1-s)q \tag{5}$$

$$P(\text{G1: } aa \text{ and } aa) \approx 1 - 4(1-s)q \tag{6}$$

Then, in the next generation G2, the probability of $Aa$ genotypes is:

$$P_{G2}(\text{inherited } Aa) = P(\text{G1: } Aa \text{ and } aa) \times \frac{1}{2} \approx 2(1-s)q \tag{7}$$

$$P_{G2}(\textit{de novo } Aa) \approx 2\nu \tag{8}$$

Assume equilibrium, in G2 we have

$$q = P_{G2}(Aa)/2 = (P_{G2}(\text{inherited } Aa) + P_{G2}(\text{de novo } Aa))/2 \tag{9}$$

therefore,

$$q \approx \frac{2(1-s)q}{2} + \frac{2\nu}{2} = \frac{\nu}{s} \tag{10}$$

Replacing $q$ with $\nu/s$, we rewrite Equation 3 and 4 (the genotypic composition of G1 individuals that would be parents) into:

$$\begin{aligned} P_{G1}(Aa) &\approx \frac{2(1-s)\nu}{s} \\ P_{G1}(aa) &\approx 1 - \frac{2(1-s)\nu}{s} \end{aligned} \tag{11}$$

and the genotypic composition of G2 pre-selection individuals:

$$\begin{aligned} P_{G2}(\text{inherited } Aa) &\approx \frac{2\nu(1-s)}{s} \\ P_{G2}(\textit{de novo } Aa) &\approx 2\nu \\ P(aa) &\approx 1 - \frac{2\nu}{s} \end{aligned} \tag{12}$$

Given penetrance $p$, we get Phenotype ∩ Genotype of G2 individuals with $Aa$ genotypes in 4 categories:

$$\begin{aligned} P_{G2}(\text{Affected} \cap \text{inherited } Aa) &= \frac{2p\nu(1-s)}{s} \\ P_{G2}(\text{Affected} \cap \text{de novo } Aa) &= 2p\nu \\ P_{G2}(\text{Unaffected} \cap \text{inherited } Aa) &= \frac{2(1-p)\nu(1-s)}{s} \\ P_{G2}(\text{Unaffected} \cap \text{de novo } Aa) &= 2(1-p)\nu \end{aligned} \tag{13}$$

From these equations, we can deduce the *de novo* mutation fraction ($\boldsymbol{F}$) in G2 $Aa$ genotypes among individuals with or without the clinical condition:

$$F_{all} = \frac{P_{G2}(\text{de novo } Aa)}{P_{G2}(Aa)} = s \tag{14}$$

$$F_{\text{affected}} = \frac{P_{G2}(\text{de novo } Aa|\text{Affected})}{P_{G2}(Aa|\text{Affected})} = s \tag{15}$$

$$F_{\text{unaffected}} = \frac{P_{G2}(\text{de novo } Aa|\text{Unaffected})}{P_{G2}(Aa|\text{Unaffected})} = s \tag{16}$$

Thus, $\boldsymbol{F = s}$ applies to random samples in the new generation G2, regardless of their condition.

However, our cohort is not randomly sampled from a population. The autism cohorts were ascertained this way: first, affected individuals were (near-) randomly enrolled in the study as the proband cases; then, a much smaller number of affected siblings from the same families were enrolled as additional cases, and a subset of unaffected siblings from the same families as controls (instead of enrolling unaffected children from population). Here we prove that the de novo fraction is lower in the affected and unaffected siblings.

$$\begin{aligned} P(\text{G2 is Affected}|\text{G1: } Aa \text{ and } aa) &= \frac{p}{2} \\ P(\text{G2 is Affected}|\text{G1: } aa \text{ and } aa) &= 2pv \end{aligned} \tag{17}$$

Using Bayes rule, with Equation 5, 6 and 10, we have the genotype frequency of G1 individuals that are parents of an affected G2 individual:

$$\begin{aligned} P(\text{G1: } Aa \text{ and } aa|\text{ one affected individual in G2}) &= \frac{1-s}{1-4(1-s)v} \approx 1-\text{s} \\ P(\text{G1: } aa \text{ and } aa|\text{ one affected individual in G2}) &\approx \text{s} \end{aligned} \tag{18}$$

This approximation holds when s is a lot greater than $4v$, which is plausible as we are assuming very rare variants ($q \ll 1/4$).

Finally, the probabilities of $Aa$ genotypes in a second child of the ascertained families:

$$P(\text{2nd individual in G2: de novo } Aa|\text{ one affected individual in G2}) = 2v \tag{19}$$

$$P(\text{2nd individual in G2: inherited } Aa|\text{one affected individual in G2}) = \frac{1-s}{2} \tag{20}$$

Therefore, for variants associated with the condition, the fraction of *de novo* carried by a second individual from the ascertained families is:

$$F = \frac{P(\text{2nd individual: de novo } Aa|\text{ one affected individual})}{P(\text{2nd individual: } Aa\ |\text{one affected individual})} = \frac{4\nu}{4\nu + 1 - s} \quad (21)$$

With Equation 21, $\boldsymbol{F}$ is always smaller than $\boldsymbol{s}$ when $\boldsymbol{s}$ is greater than $4\nu$. We note $\nu$ is about $10^{-7}$ to $10^{-8}$ in human. Therefore, $\boldsymbol{F}$ is close to 0 (in same magnitude as $\nu$) for $\boldsymbol{s}$ in the range of interest of this study ($10^{-4}$ to 1), except when $\boldsymbol{s}$ is very close to 1. In the autism cohorts, most (>90%) of the cases were the ascertained probands based on which the families were enrolled in the studies, and all of the controls are unaffected siblings conditioned on one affected child in the families. Therefore, among variants that are associated with autism, $\boldsymbol{F}$ will be close to 0 in controls and be below but close to $\boldsymbol{s}$ in cases.

Empirically, a substantial fraction of variants under strong selection are associated with autism and neurodevelopmental disorders (see Supplementary Figure 13 and previous publications using metrics correlated with selection strength of protein truncating variants [4,5]), and that the chance of being associated positively correlated with the estimated selection coefficient. $\boldsymbol{F}$ of these variants is close to 0 in controls. Therefore, among all variants under strong selection, $\boldsymbol{F}$ in unaffected siblings (i.e. controls) is smaller than $\boldsymbol{s}$, leading to deviation from the diagonal of $\boldsymbol{F}$ vs $\boldsymbol{s}$ curve, and the deviation is more pronounced with greater $\boldsymbol{s}$ values (Figure 4b). In cases, the deviation is minor since most of the cases were probands with near random ascertainment.

#### MisFit performance compared with EVE

We compared our model with several popular computational methods. EVE[6] is also a state-of-art methods, which is an Bayesian variational autoencoder using multiple sequence alignments as inputs. As EVE only generates scores for 3,219 known disease genes with good MSA, we also limit the deep mutational scan data to a subset of 14 genes with all prediction scores available for fair comparison (Supplementary Fig. 19). Generally, MisFit_D has a similar performance with EVE. Here we used the EVE-predicted posterior probability of damaging component from their mixture model, which is already adjusted specifically for each gene.

In *de novo* variants analysis, variants without EVE annotations are regarded as least damaging. This largely impairs sensitivity, but the top risk variants are less affected as they are likely to come from known disease associated genes with good coverage of EVE.

#### MisFit model on predicting ClinVar variants

We also analyzed the concordance with ClinVar labelled variants. ClinVar variants were processed from the version of Dec 2022. Variants with 'criteria provided' (1 review star) were collected, and the ones with conflicting labels or annotated as 'variant of uncertain significance' were removed. Pathogenic / likely-pathogenic variants are regarded as positive and benign / likely-benign variants are regarded as negative. To eliminate the gene-level bias, we selected genes with at least one positive label and one negative label, and down-sample the variants to make equal number of positive and negative label in each gene. Finally, this gives out 33,046 variants in 3,246 genes. While AlphaMissense has a best performance in this analysis, MisFit_D is also reasonably good (Supplementary Fig. 20).

Although this analysis is informative in clinical applications, we still lack totally independent data for testing. Supervised methods are usually trained on these labels from ClinVar, or from similarly curated databases. Additionally, some methods (such as CADD and increasingly REVEL) are commonly used as one of the criteria for annotating pathogenicity in ClinVar.

#### Point estimate for selection coefficient

Under the simplest Poisson assumption, where $m \sim Pois(nv/s)$, we can easily use $s_{MLE} = nv/m$ as a point estimate by maximum likelihood estimation. However, as $s$ is in the denominator of the Poisson mean parameter, $s_{MLE}$ is naturally a biased estimator of $s$. We prove it here.

$$E(s_{MLE}) = E\left(\frac{nv}{m}\right) = nvE\left(\frac{1}{m}\right) \geq \frac{nv}{E(m)} = \frac{nv}{nv/s} = s$$

Here, $E\left(\frac{1}{m}\right) \geq \frac{1}{E(m)}$ , because $f(x) = 1/x$ is a convex function, and thus $E\big(f(x)\big) \geq f(E(x))$ (Jensen's inequality).

When considering a more realistic population genetics model, the situation can be more complicated, but $s_{MLE} = argmax(p(data|s))$ is still biased because the relationship as a denominator holds. Adding a prior distribution may alter the estimation. Although those Bayes approaches enable describing a whole posterior distribution of $s$, we usually still need to derive a point estimate for easier downstream analysis. As there are no standard criteria of doing this, several previous studies[3,7,8] used posterior mean as point estimate, defined as $E(s|data)$. In this study, because every $s$ in the model is transformed in logit scale with $s' = \log\left(\frac{s}{1-s}\right)$, we directly used posterior mean in the logit scale $E(s'|data)$, and transformed it back to original scale, which gives $sigmoid(E(s'|data))$. This value is different with $E(s|data)$ when limited data are provided, but could be potentially less biased. (Supplementary Fig. 21)

##### Improve estimation of selection by using genomes from different populations

We showed that adding sequencing samples from the same population does not help with estimating small select coefficient, but adding samples from another population improves the estimation. Assuming the second population evolves totally independently from the first population, such improvement is comparable to doubling the integrated number of variants with same selection coefficient. If the two populations split very recently, the allele frequencies are highly correlated and provide limited additional information, and the result should be the same as adding samples from the same population. We simulated the allele frequencies in two hypothetic populations assuming different length of independent evolution (Supplementary Fig. 22). 2,000 generations (approximating split time of Europeans and Africans) is already long to give out less correlated data, except for very common variants with allele frequencies larger than 0.1.

### Supplementary Figures

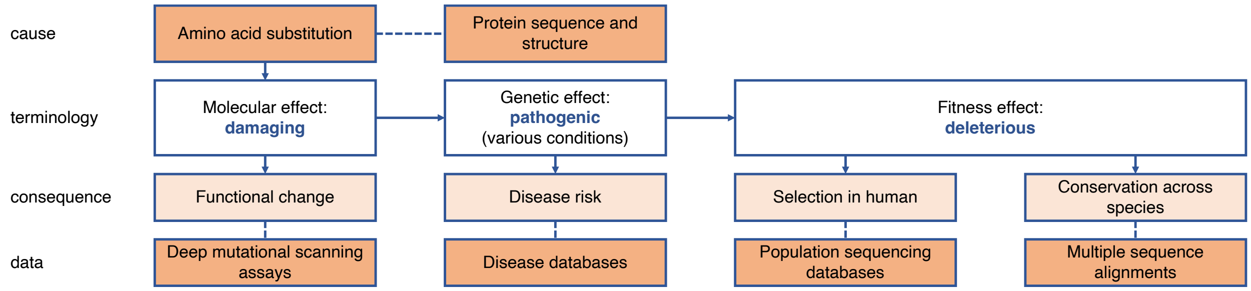

#### Supplementary Fig 1 Missense variant effect from different aspects

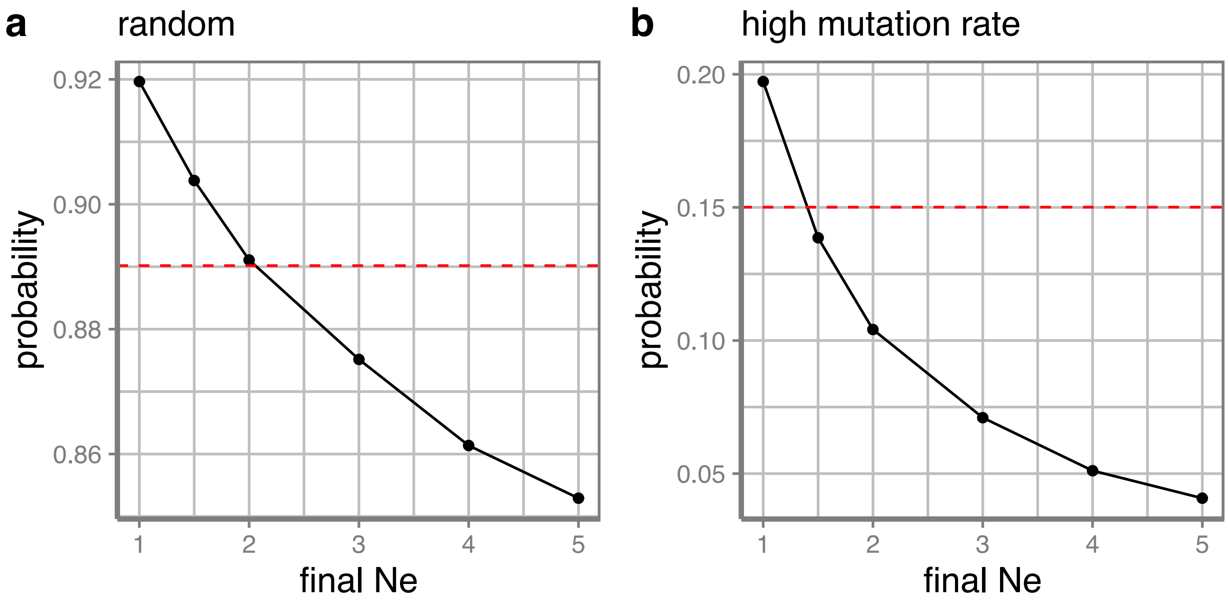

**Supplementary Fig 2 Adjusted European final effective population size in simulation**

Synonymous variants are simulated based on European effective population size history with different final population size in the latest generation. Probabilities of zero-count allele in simulation (black) and in UKBB plus gnomAD NFE samples (red) are shown. **a** synonymous variants are randomly selected with an average mutation rate of 1e-8. **b** Only C-to-T synonymous variants in CpG sites with mutation rate larger than 1e-7 are selected.

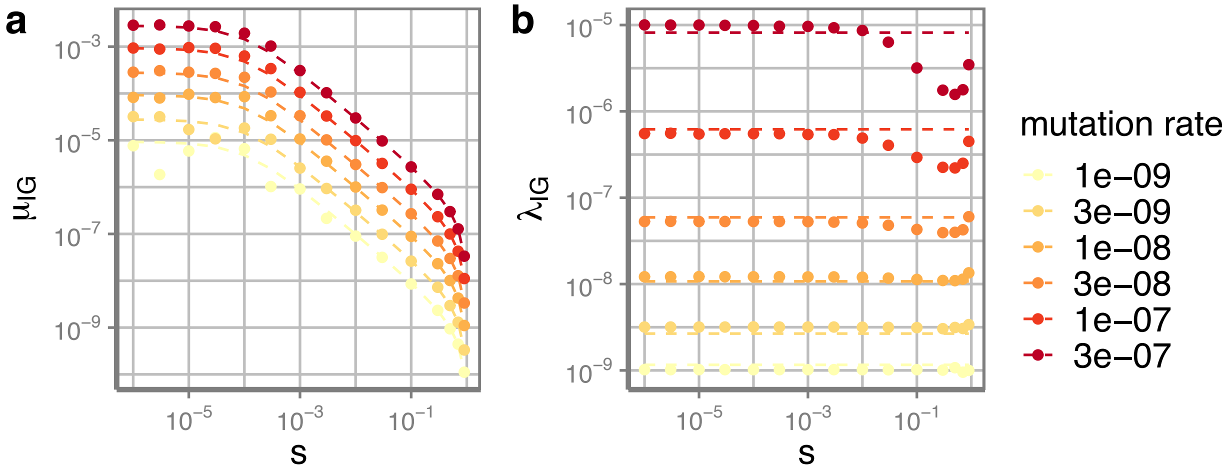

**Supplementary Fig 3 Parameters of Poisson-Inverse-Gaussian model**

$\mu_{IG}$ and $\lambda_{IG}$ are mean and shape parameters for an Inverse Gaussian distribution for modeling population allele frequency. Dots are the best fits for each simulation condition, while dashed lines are from optimized functions of mutation rate and selection coefficient.

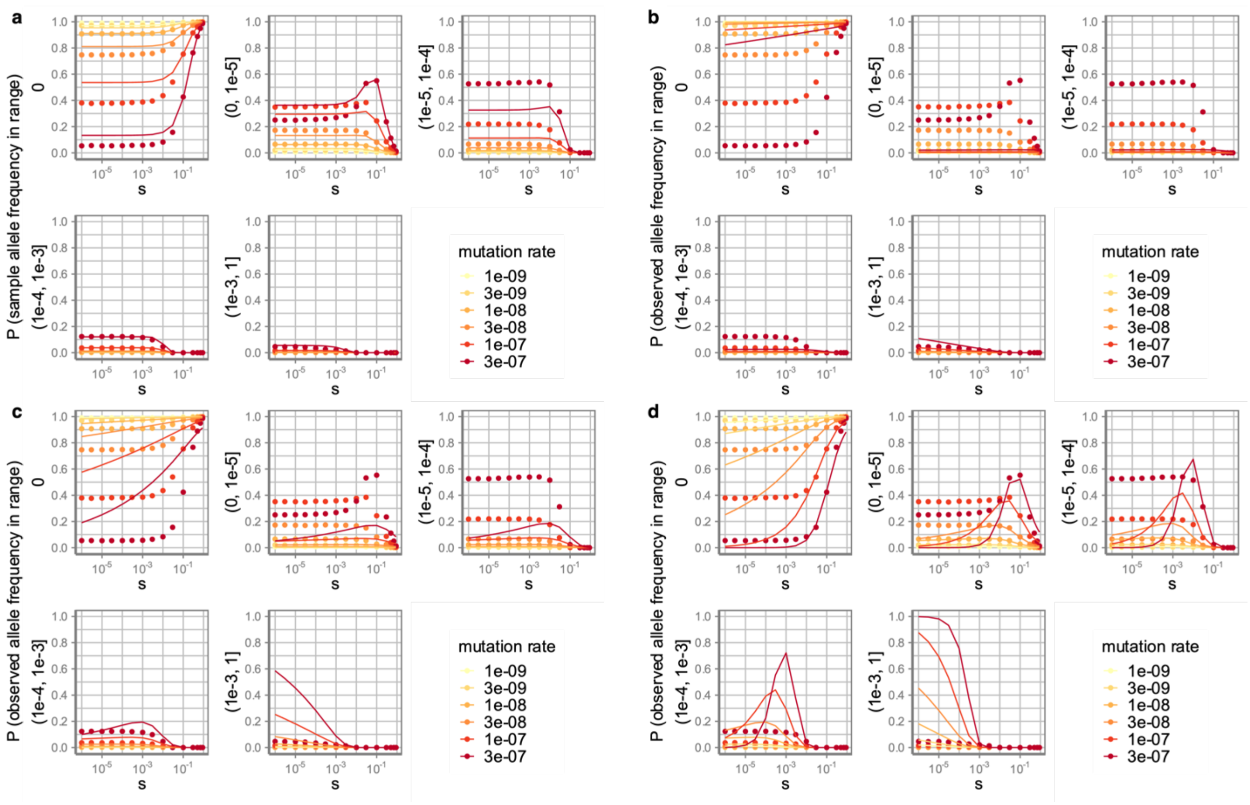

**Supplementary Fig 4 Distribution of sample allele frequency under different population genetics model**

**a** PIG model used in MisFit **b-d** Negative Binomial distribution with effective population size of **b** 10,000 **c** 100,000 **d** 1,000,000. Sample size is 200K diploid genomes. Y-axis shows a probability mass cumulated for allele frequency within a range that is annotated vertically next to the y axis.

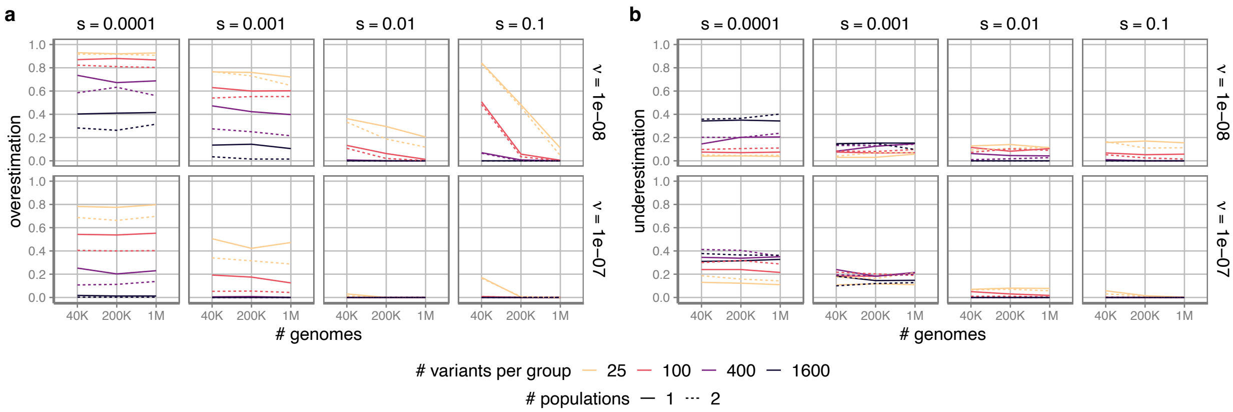

#### Supplementary Fig 5 Evaluation of MLE estimation of *s*

Probabilities of **a** overestimation **b** underestimation in 400 replications for each simulation condition are shown. Here *s* is a categorical variable of [0.00001, 0.0001, 0.001, 0.01, 0.1, 1]. Each group contains a certain number of variants (x-axis) with same *s*. Solid lines are samples from a single population, while dashed lines are samples from two populations (half of the indicated number for each population).

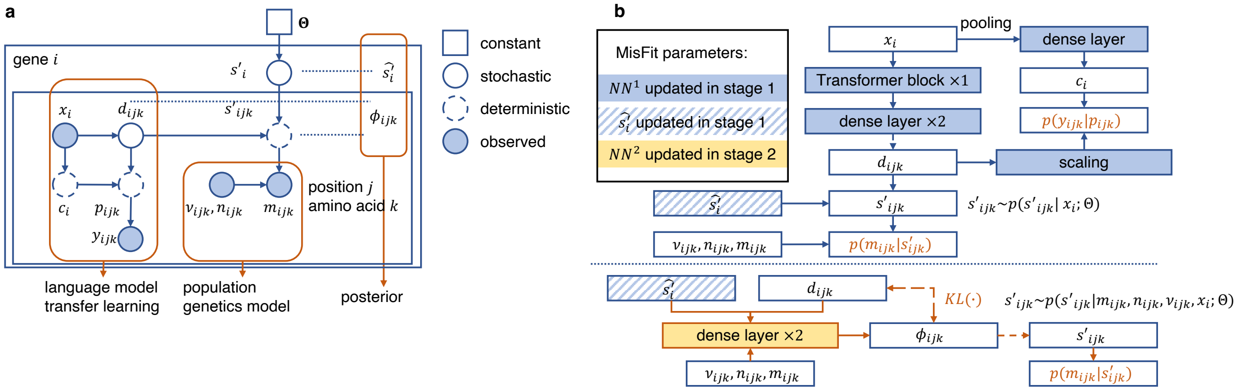

$x_i$: protein embedding from sequence; $d_{ijk}$: degree of damage to protein; $p_{ijk}$: probability of homologous variants; $y_{ijk}$: indicator of homologous variants; $c_i$: gene-level constraint scaler across species; $s'_{ijk}$: variant heterozygous selection coefficient (logit scale); $s'_i$: gene-level selection coefficient (logits scale); Θ: global prior; $v_{ijk}$: mutation rate; $n_{ijk}$: allele number in samples; $m_{ijk}$: allele count in samples; $\hat{s'_i}$: maximize a posteriori gene-level selection coefficient (logit scale); $\phi_{ijk}$: variational inference parameters for posterior $p(s'_{ijk}|m_{ijk}, n_{ijk}, v_{ijk}, x_i; \Theta)$

#### Supplementary Fig 6 Overview of MisFit model

**a** MisFit model in view of a probabilistic graphical model. **b** Full structure of MisFit model and training stages. Loss related terms are colored.

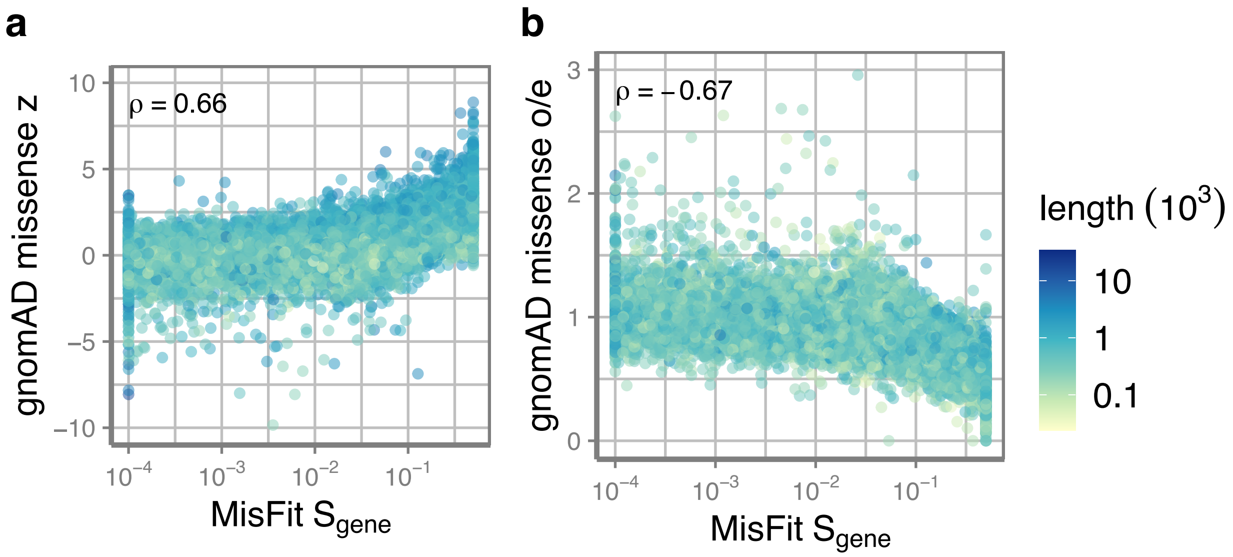

**Supplementary Fig 7 MisFit estimated gene-level missense selection correlates with gnomAD missense z score or o/e.**
Spearman correlation coefficient is annotated.

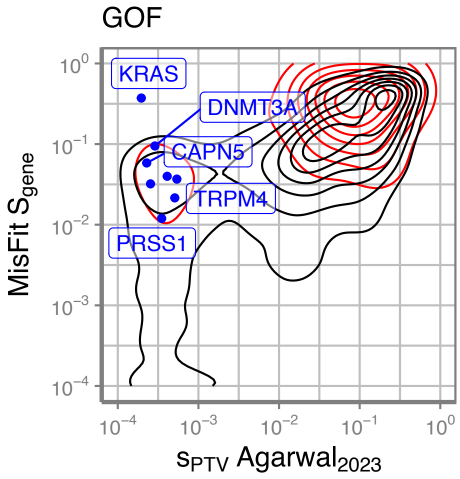

**Supplementary Fig 8 Distribution of gene-level selection for genes with known gain-of-function mechanism.**

Genes with MisFit $S_{gene} > 0.01$ but $S_{PTV} < 0.001$ (Agarwal 2023) are labeled.

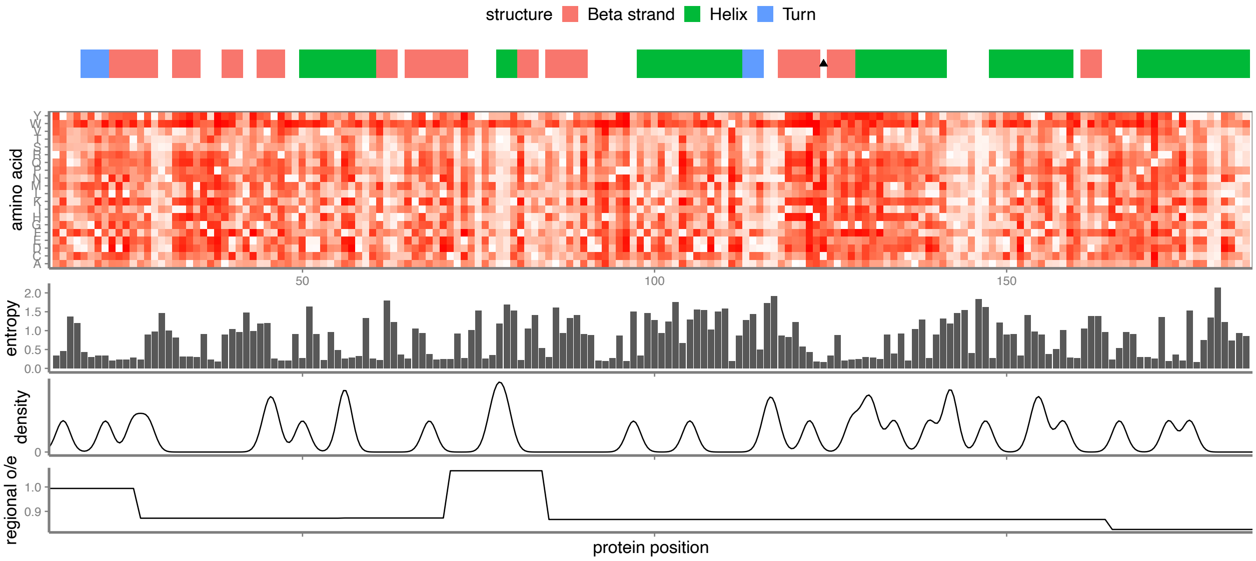

##### Supplementary Fig 9 Predicted selection coefficient for each amino acid substitution in PTEN

Secondary structures from UniProt are shown at the top. Darker color in heatmap corresponds to relatively larger selection coefficient. Entropy is calculated by amino acid distribution across Ensembl homologues, lower the value means more conservation. Missense variant density combines UKBB and gnomAD NFE populations. Regional o/e is extracted from gnomAD-3 1kb window.

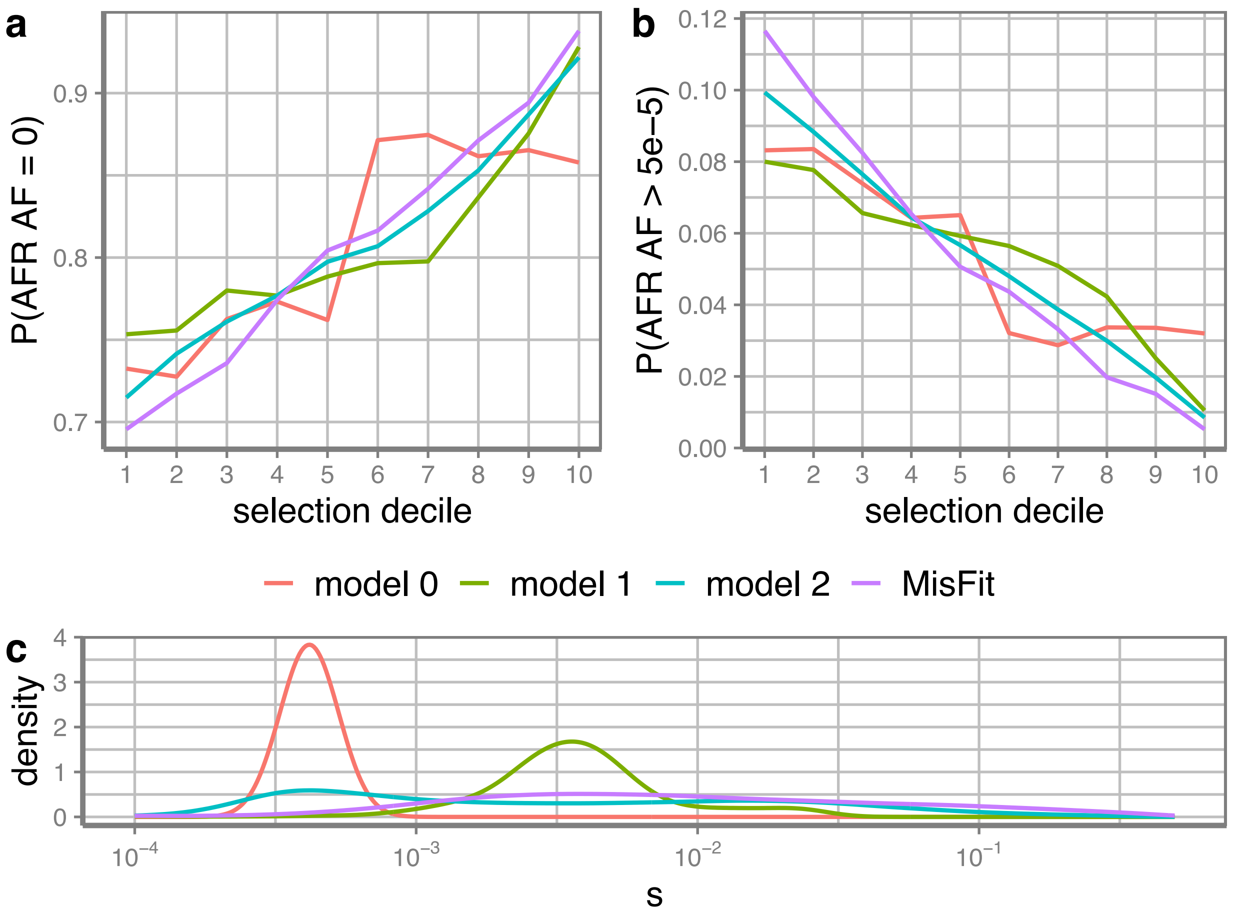

**Supplementary Fig 10 MisFit estimated selection coefficient s predict allele frequency in a second population better than baseline models**

Variants of high mutation rate (>1e-7) with sample AF < 5e-6 in training set (UKBB + gnomAD NFE) are selected for analysis. Variants are separated into 10 groups by estimated s in each model. The proportions of variants with **a** gnomAD AFR sample AF = 0 or **b** gnomAD AFR sample AF > 5e-5 are shown. **c** Distribution of estimated s of these variants. Model 0: baseline model learned only from NFE allele number, NFE allele count, and mutation rate; model 1: model 0 + gene-level selection; model 2: model 1 + ESM-2 zero shot as $d$; MisFit: model 1 + ESM-2 embeddings as $d$.

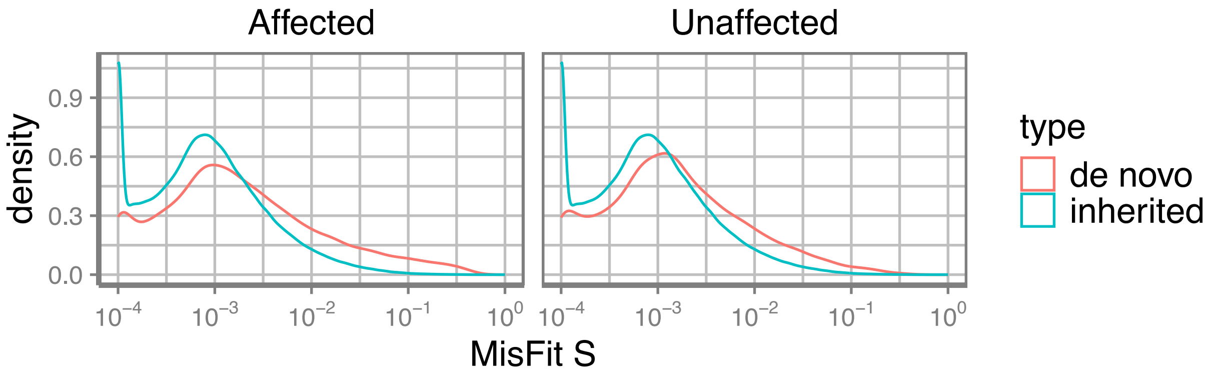

**Supplementary Fig 11 Distribution of inherited or de novo missense variants in autism dataset**

Two-sided Kolmogorov-Smirnov tests between de novo variants and inherited variants give out $p$-value in both affected and unaffected lower than numerical precision.

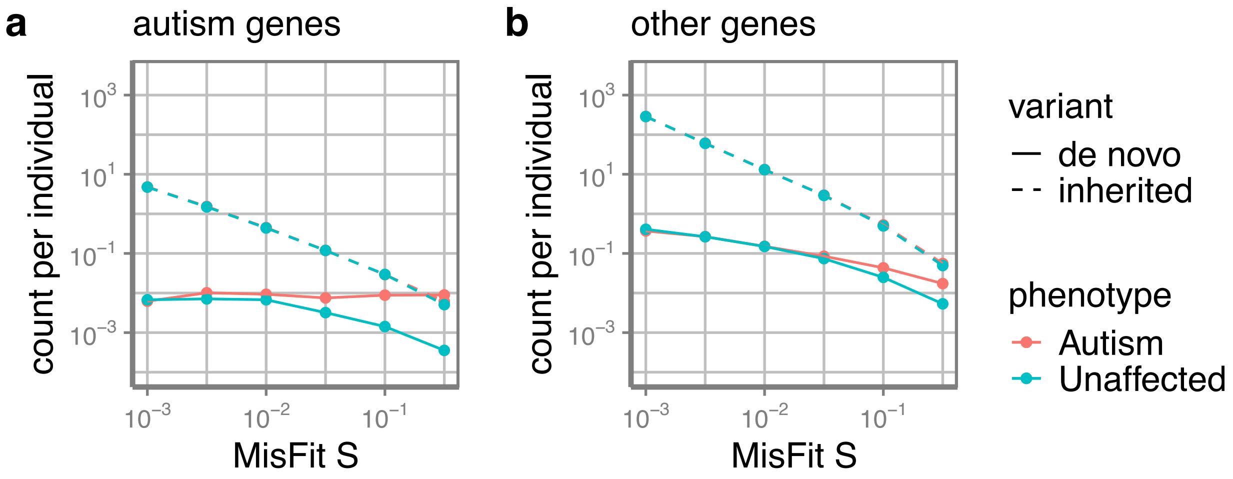

**Supplementary Fig 12 Count of de novo or inherited variants binned by of MisFit_S in different gene sets**

Dashed line indicates the inherited, solid line indicates the *de novo*. **a** known 162 autism genes from SPARK. **b** other genes.

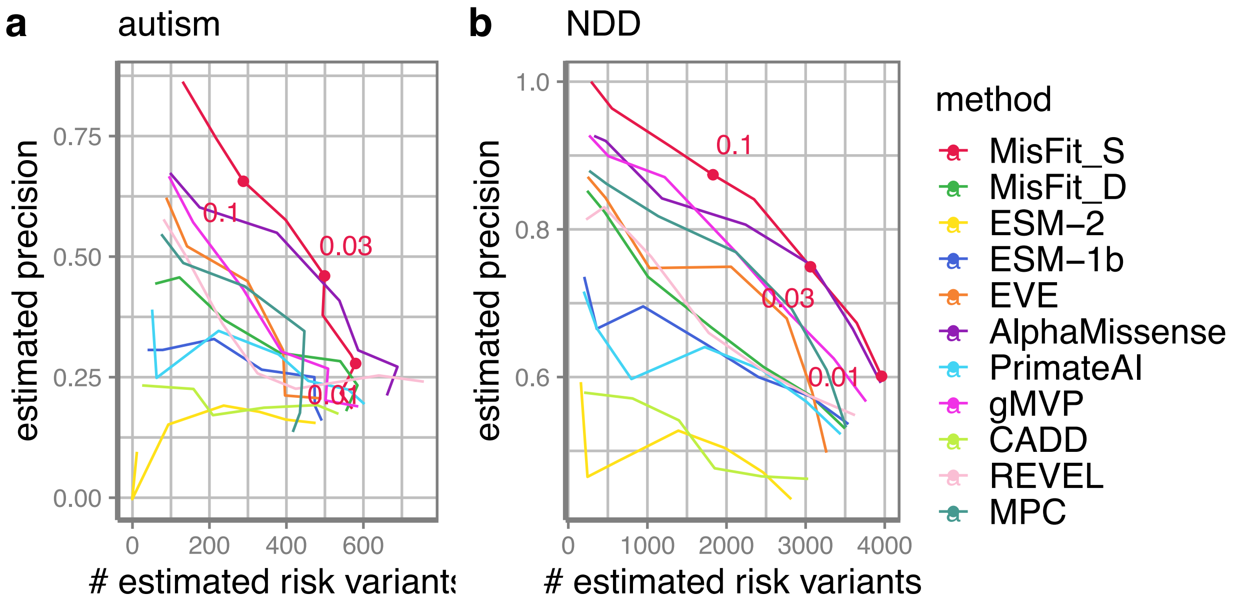

**Supplementary Fig 13 Precision-recall-proxy curve for de novo missense variants**

Thresholds of MisFit_S are annotated.

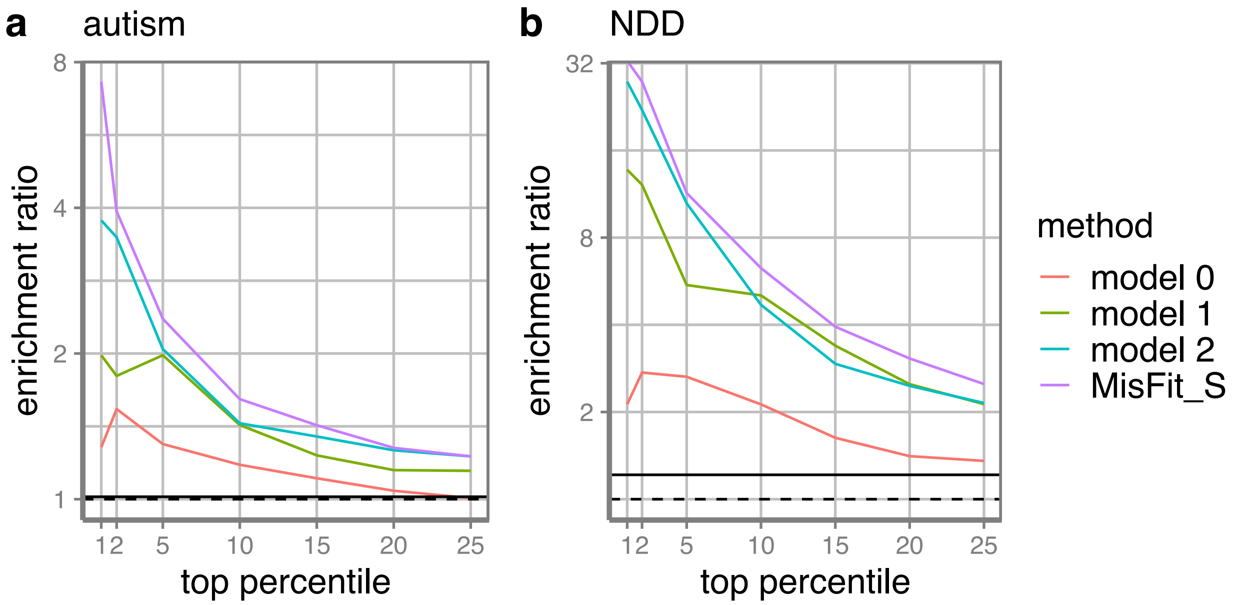

**Supplementary Fig 14 Enrichment of de novo variants in baseline models**

Model 0: baseline model learned only from NFE allele number, NFE allele count, and mutation rate; model 1: model 0 + gene-level selection; model 2: model 1 + ESM-2 zero shot as $d$; MisFit: model 1 + ESM-2 embeddings inferred $d$.

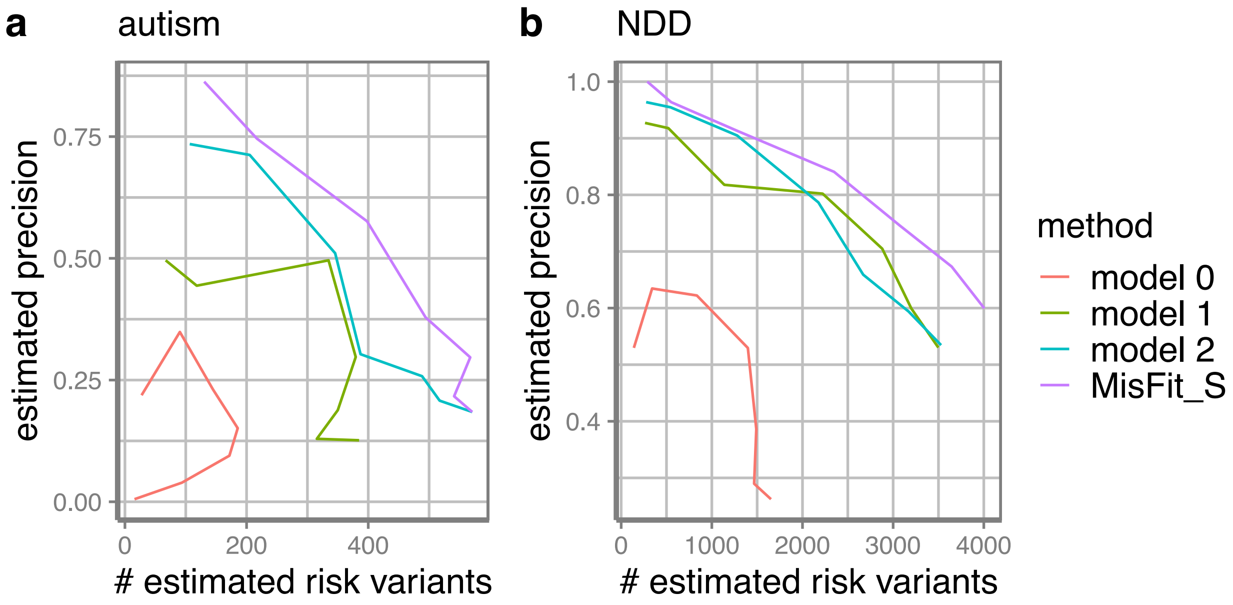

**Supplementary Fig 15 Precision-recall-proxy curve of de novo variants in baseline models**

Model 0: baseline model learned only from NFE allele number, NFE allele count, and mutation rate; model 1: model 0 + gene-level selection; model 2: model 1 + ESM-2 zero shot as $d$; MisFit: model 1 + ESM-2 embeddings inferred $d$.

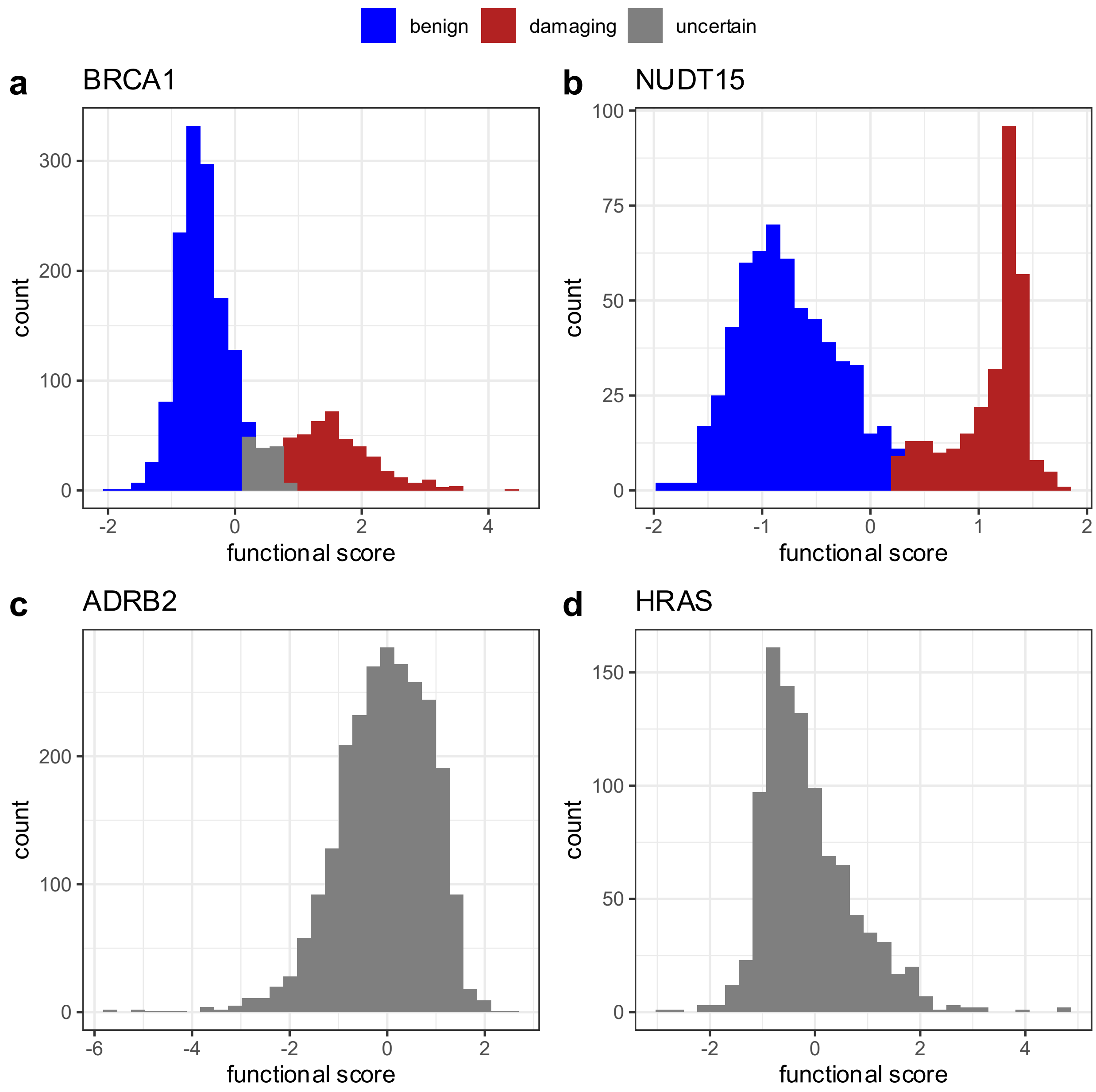

**Supplementary Fig 16 Various functional score distributions in different deep mutational scanning experiments**

**a-b** Examples of bimodal distribution with originally annotated labels. **c-d** Examples of unimodal distribution.

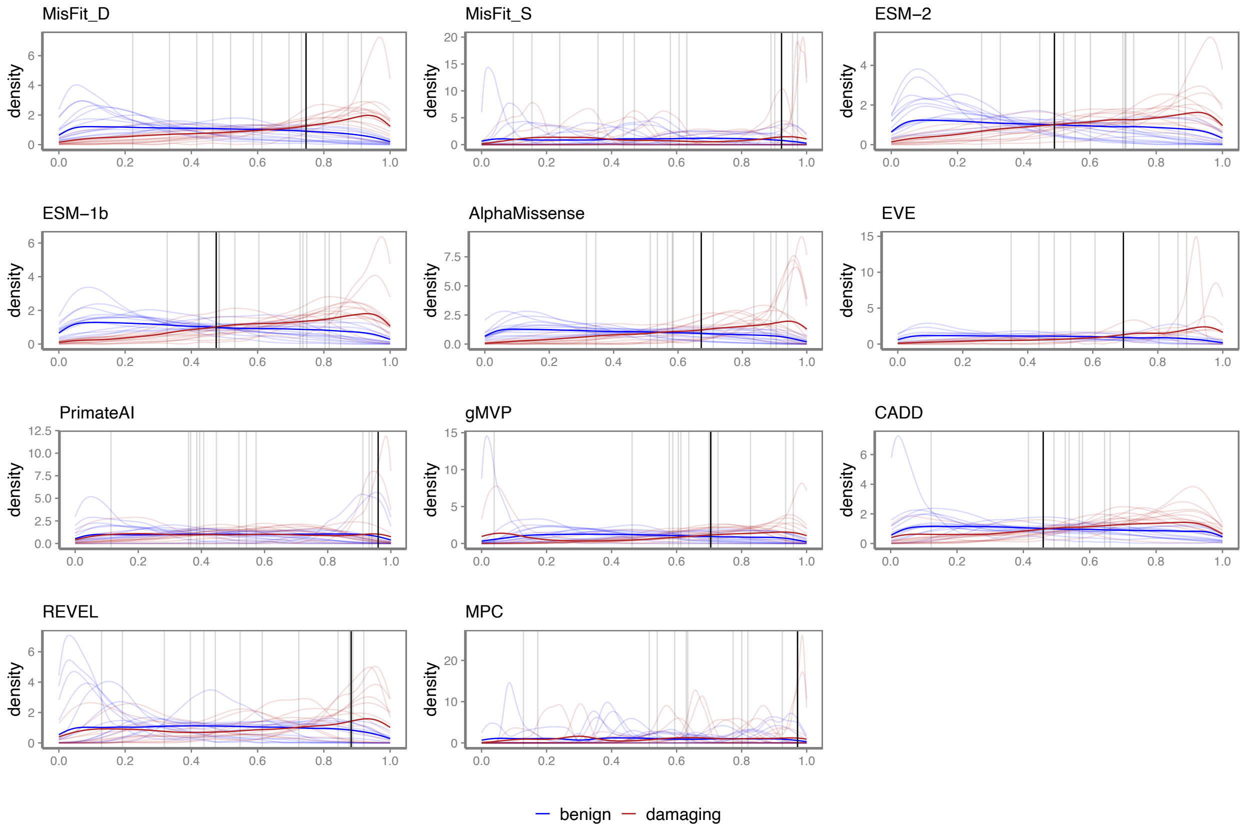

#### Supplementary Fig 17 Distribution of scores (normalized by rank) across genes

The red and blue curves show the distribution of damaging and benign variants, respectively. Dark curves are the distribution of scores in the combined data, while light curves show each gene separately. The black lines are the optimal threshold achieving highest MCC in the combined dataset, while the grey lines are the optimal threshold for each gene.

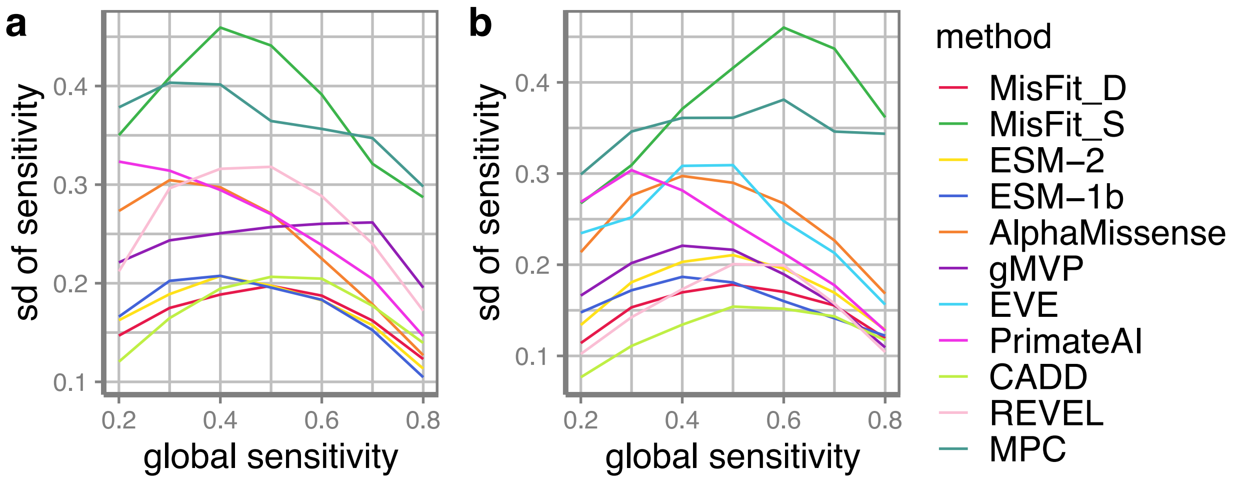

**Supplementary Fig 18 Consistency of sensitivity across genes**

Thresholds are set to achieve certain global sensitivity (x-axis) in the combined data. Sensitivities in different genes are then evaluated. Y-axis shows the standard deviation. **a** in all genes; **b** in EVE genes.

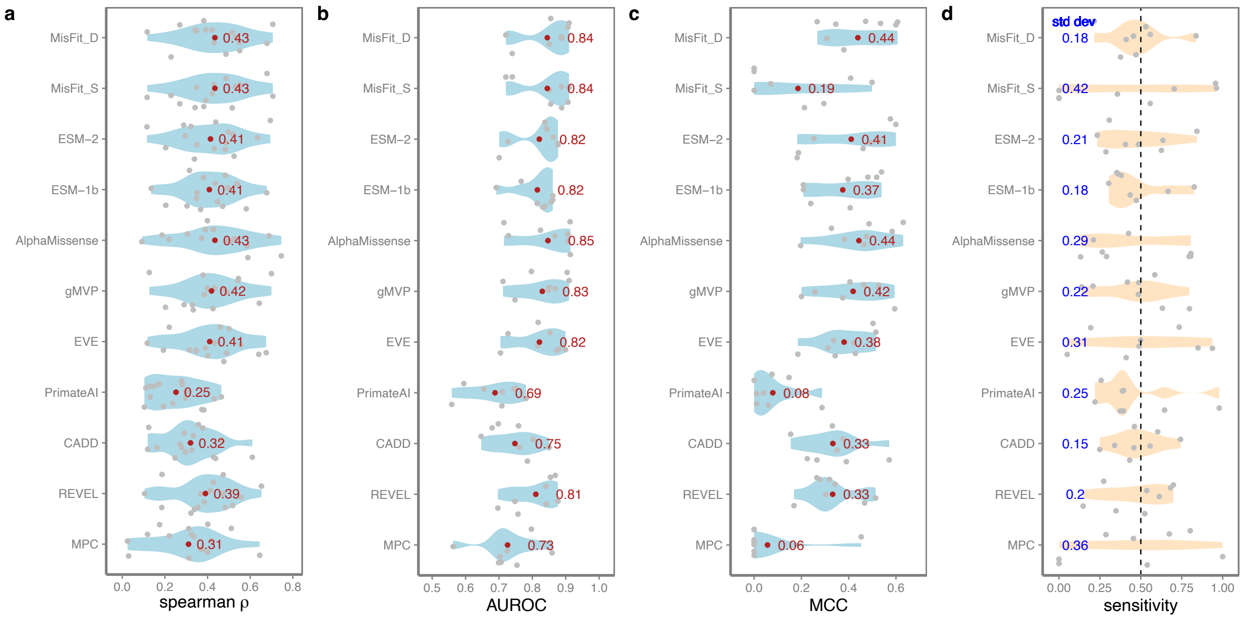

#### Supplementary Fig 19 Performance in predicting damaging variants in deep mutational scanning assays and cross-gene consistency

A subset of genes from **Fig. 7** with all prediction scores available are used for analysis. **a** Spearman correlation coefficient of predicted scores with functional scores from deep mutational assays. Mean is annotated in red. **b** AUROC of predicting confidently labeled damaging or benign variants in deep mutational assays. Mean is annotated in red. **c** MCC in each gene with a global threshold that achieves best MCC in the combined dataset. Mean is annotated in red. **d** Sensitivity in different genes when setting a threshold to achieving a global sensitivity of 0.5 (dashed) in the combined dataset. Standard deviation is annotated in blue. For **b-d**, different assays of same gene are combined so that variants with a damaging label in any of the assays will be regarded as damaging.

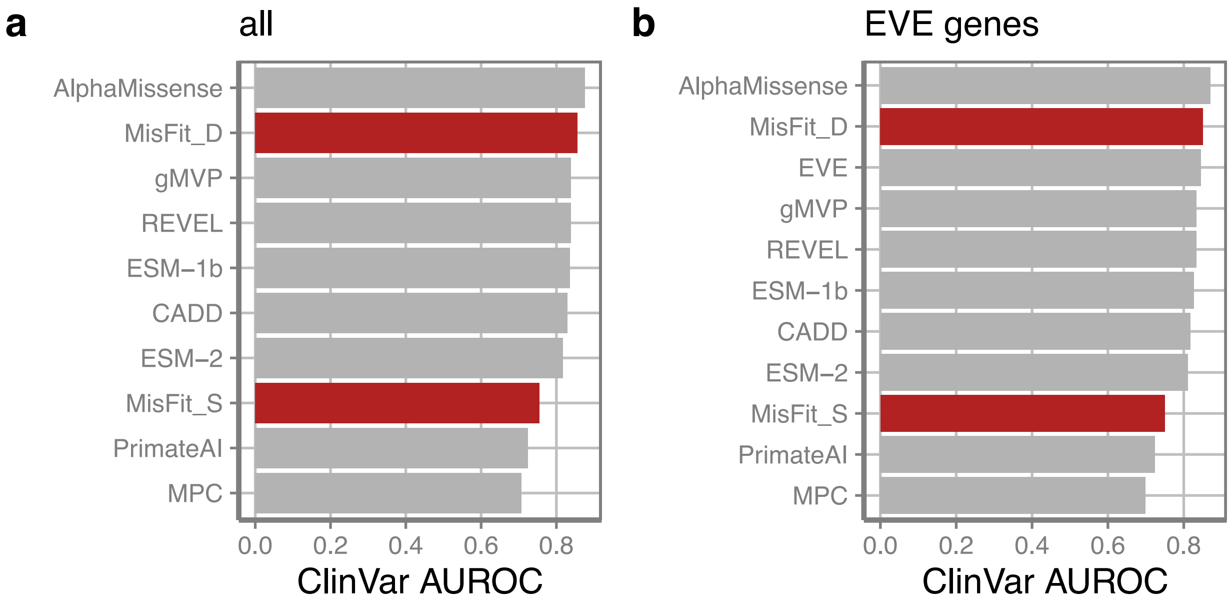

**Supplementary Fig 20 AUROC in predicting ClinVar balanced pathogenic / benign variants**

**a** 33,046 variants in 3,246 genes. **b** 26,171 variants in 2,229 genes with all prediction scores available.

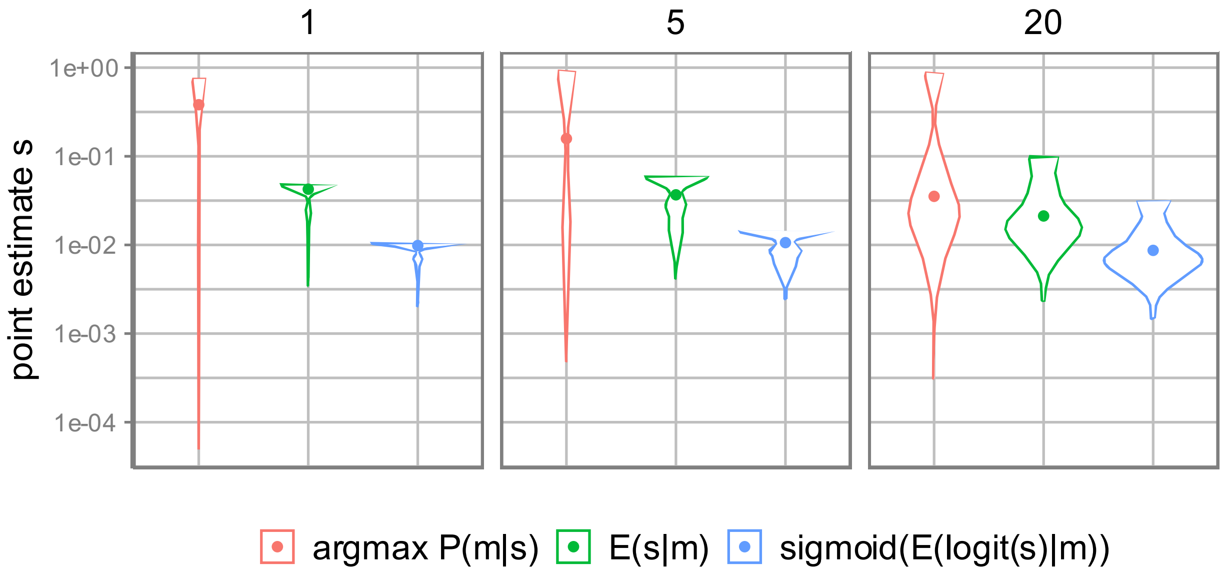

**Supplementary Fig 21 Bias of point estimate of heterozygous selection coefficient**

The number on top shows the aggregated number of variants per group, and y-axis shows the distribution of $s$ estimated for each group in 100 replicates. The simulation condition is $s = 0.01$, so the mean of point estimate (dot) closer to this value meaning the estimator is less biased. Here mutation rate is 1e-8 and sample size is 200K.

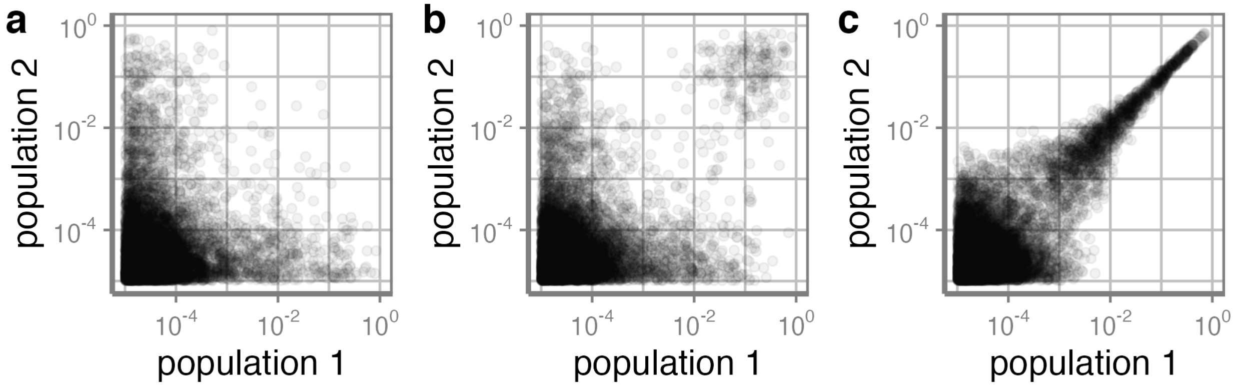

**Supplementary Fig 22 Correlation of population allele frequency in two populations**

The two simulated populations split **a**. 10,000 generations ago **b** 2,000 generations ago **c** 200 generations ago. Mutation rate is 1e-7 and heterozygous selection coefficient equals 1e-4.

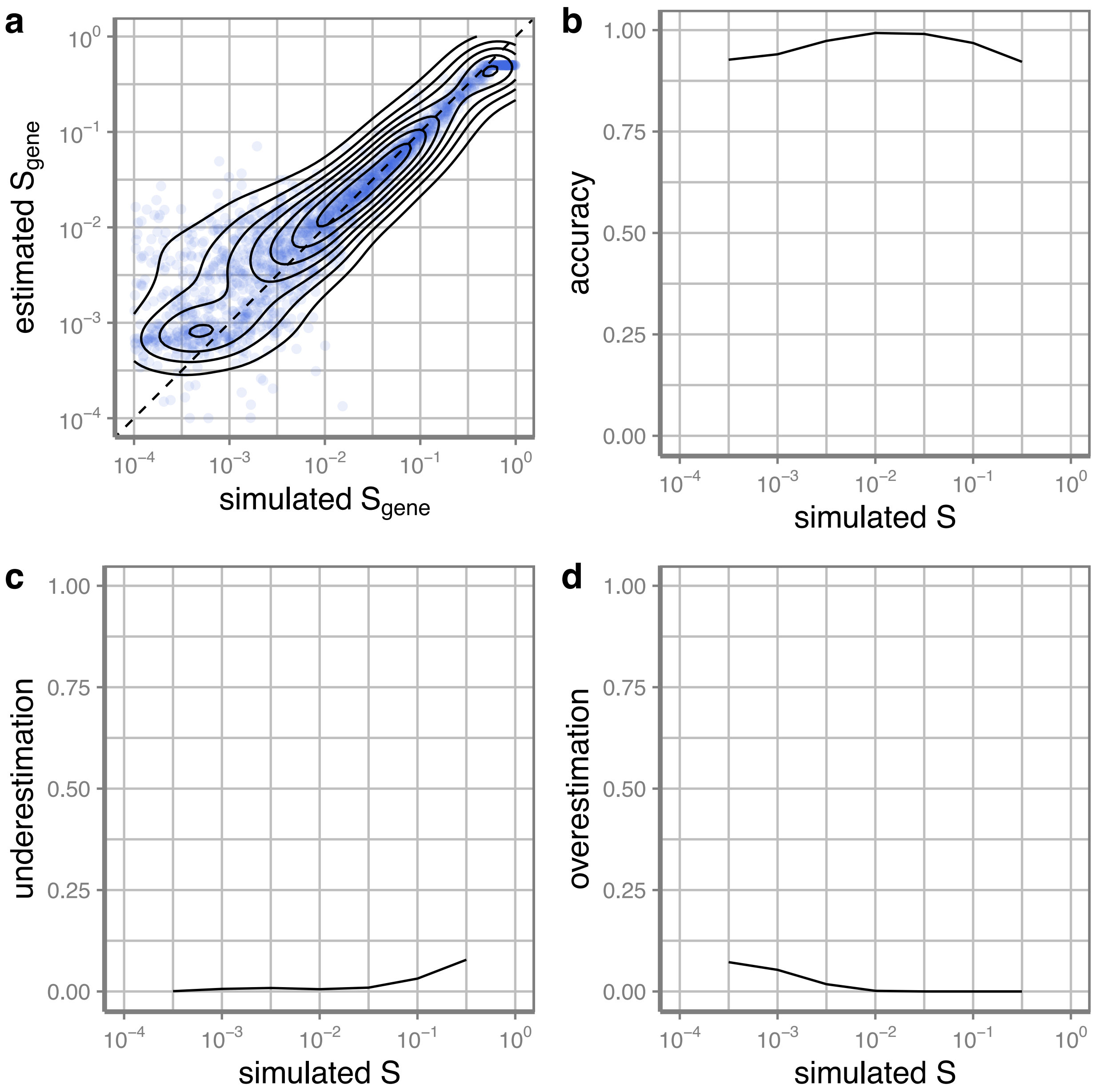

**Supplementary Fig 23. Estimated model performance in simulation.** Selection coefficients and allele frequencies of variants in 2000 genes are simulated based on MisFit probabilistic model assumptions. **a**. Estimated $S_{gene}$ (prior of max $S$ in a gene) versus simulated. **b-d** Accuracy/underestimation/overestimation rate of variant selection coefficient. An accurate estimation is defined as $(10^{-0.5} \times \text{simulated } S) \leq$ estimated $S \leq (10^{0.5} \times \text{ simulated } S)$.

### Supplementary Tables

#### Supplementary Table 1

MisFit estimated gene-level selection for 830 genes with known disease mechanisms.

#### Supplementary Table 2

Genetic variants data used in analysis.

#### Supplementary Table 3

Deep mutational scanning experiments used in analysis and Spearman correlation coefficient with computational methods.

#### Supplementary Table 4

A subset of deep mutational scanning experiments that have bimodal distribution of functional scores used in analysis, and AUROC of computational methods.

#### Supplementary Table 5

MisFit estimated gene-level selection (including missense and protein-truncating variants) for all genes trained in model.
